# Meteorological Drivers of Influenza A and B Positivity in a Subtropical Chinese City: A Six-Year Surveillance Study Integrating Distributed Lag Non-Linear Models and Deep Learning

**DOI:** 10.1101/2025.07.13.25331478

**Authors:** Long Xie, Meng-Jie Zhang, Jin-Lin Tan, Yi-Xin Ling, Zhe-Qiang Xue, Zhi-Shen Wu, Jun-Ju Huang, Jian-Ling Chen, Ze-Fan Ruan, Jing Qian, Hai-Yong Pan, Xiao Han, Sheng Xiong, Long-Mei Ling, Xi-Wen Jiang

**Affiliations:** Translational & Clinical Research Institute, Faculty of Medical Sciences, Newcastle University, Newcastle upon Tyne, UK; Department of Microbiology Laboratory, Putian Centre for Disease Control and Prevention, Putian, China; Research Institute, DAAN Gene Co., Ltd, Guangzhou, China; The Medicine and Biological Engineering Technology Research Centre of the Ministry of Health, Guangzhou, China; College of Biological Science and Engineering, Fuzhou University, Fuzhou, China; Department of Infection, Mengchao Hepatobiliary Hospital of Fujian Medical University, Fuzhou, China; College of Life Science and Technology, Jinan University, Guangzhou, China; School of Life Sciences and Biopharmaceutics, Guangdong Pharmaceutical University, Guangzhou, China

**Keywords:** Influenza Positivity Rate, Infectious Disease Dynamics, Climate-Sensitive Digital Surveillance, Surveillance Bias, Public Health Forecasting Framework, Subtropical Epidemiology

## Abstract

Influenza transmission in subtropical regions is shaped by complex, non-linear meteorological conditions, while surveillance-based forecasting has been further complicated by fluctuating testing intensity and non-pharmaceutical interventions during the COVID-19 period. Existing approaches often either characterize lagged environmental associations without forecasting capacity or apply deep learning without clear epidemiological interpretability. Using six years (2018-2023) of multi-site influenza surveillance data from Putian, a subtropical coastal city in southeastern China, we developed a two-stage framework to characterize meteorological associations and improve short-horizon forecasting. Influenza positivity rates were used as the primary outcome to mitigate testing-related surveillance bias. Distributed lag non-linear models (DLNMs) were constructed to estimate subtype-specific, lagged associations between meteorological variables and influenza positivity, and a Bayesian-optimized long short-term memory (LSTM) network integrating meteorological, autoregressive, and socio-behavioral covariates was developed to forecast influenza A and B activity. Influenza A positivity was associated with warmer conditions, peaking at 15°C (relative risk [RR] = 3.01, 95% CI: 1.03-8.78) and narrow diurnal temperature ranges, whereas influenza B positivity was associated with colder (8°C; RR = 26.12, 95% CI: 7.79-87.61), more humid (91%; RR = 1.52, 95% CI: 1.00-2.29), and lower-radiation conditions. The LSTM achieved lower forecast error than covariate-matched baselines (multivariate Autoregressive Integrated Moving Average [ARIMA] and eXtreme Gradient Boosting [XGBoost]) for both subtypes (Mean Absolute Error [MAE] for influenza A: 0.009 vs. 0.136 [ARIMA] and 0.138 [XGBoost]; for influenza B: 0.002 vs. 0.049 [ARIMA] and 0.070 [XGBoost]; Symmetric Mean Absolute Percentage Error [SMAPE] for influenza A: 0.521 vs. 1.212 and 1.081; for influenza B: 0.484 vs. 0.810 and 0.737, respectively). Covariate-ablation and SHapley Additive exPlanations (SHAP) analyses suggested that historical positivity and testing-related variables contributed materially to forecast stability under surveillance variability. External evaluation in Sanming provided preliminary evidence of model portability. This integrated DLNM-LSTM framework offers an interpretable approach for characterizing subtype-specific meteorological associations and improving influenza forecasting in subtropical settings.

**Importance:** Influenza imposes substantial global morbidity and mortality each year, and forecasting is particularly difficult in subtropical regions, where transmission occurs year-round, meteorological drivers are non-linear and lagged, and the COVID-19 period introduced pronounced epidemiological non-stationarity. Forecasts based on raw case counts are further affected by surveillance-related bias, as counts partly reflect testing intensity and clinician vigilance rather than underlying transmission alone. Existing work has largely applied inferential and deep-learning approaches in isolation, limiting either interpretability or forecasting flexibility. We address these issues with a two-stage DLNM–LSTM framework that uses influenza positivity rates as the primary endpoint and incorporates weekly detection volumes, mask-wearing stringency indices, day-and-week and non-local population proportion as socio-behavioral covariates, in order to mitigate testing-related surveillance bias and accommodate pandemic-era non-stationarity. The framework reveals distinct, subtype-specific meteorological associations for influenza A and B, shows lower forecast errors than covariate-matched baselines, both linear autoregressive (ARIMA) and non-linear machine-learning (XGBoost), and provides preliminary evidence of portability to a second subtropical city. It offers an interpretable, climate-informed methodological foundation for future operational surveillance developments in subtropical influenza forecasting.

## Introduction

Influenza imposes a substantial global health burden, with an estimated 290,000–650,000 respiratory deaths and 3–5 million severe cases among one billion infections annually (Iuliano et al. 2018). In China alone, influenza is associated with an estimated 88,100 excess respiratory deaths per year (Li et al. 2019), with children, the elderly, and individuals with chronic comorbidities disproportionately affected. Effective preparedness therefore requires both a mechanistic understanding of the drivers of influenza activity and the operational capacity to forecast its short-term trajectory. Because influenza A and B viruses are transmitted primarily through respiratory droplets and contact (Uyeki et al. 2022), with transmission dynamics principally influenced by viral characteristics, host susceptibility, and environmental conditions (Javanian et al. 2021), meteorological variables, particularly temperature, humidity, and precipitation, represent a critical, potentially modifiable dimension of transmission (Moriyama, Hugentobler, and Iwasaki 2020).

Meteorological drivers of influenza transmission have been extensively characterized in temperate settings, where a pronounced winter peak (Ryu and Cowling 2021) is associated with low ambient temperatures (5-8°C) that stabilize aerosolized influenza virions and impair host mucociliary clearance (Moriyama, Hugentobler, and Iwasaki 2020), and with low absolute humidity (<40%) that prolongs airborne aerosol residence while compromising respiratory epithelial defenses (Peci et al. 2019; Lowen and Steel 2014; Eccles 2002; Tamerius et al. 2013). However, these temperate-zone patterns do not generalize to subtropical regions, where influenza circulates year-round, exhibits multiple annual peaks, and demonstrates meteorological coupling that differs substantially in directionality and lag structures from temperate observations (Deyle et al. 2016; Soebiyanto et al. 2015). Despite the demographic and climatic significance of subtropical regions for global influenza dynamics (Bloom-Feshbach et al. 2013; Tamerius et al. 2013), dedicated quantitative studies in subtropical Chinese urban areas, particularly in non-megacity settings that nonetheless harbor substantial populations and represent prevailing regional urbanization patterns, remain sparse. This geographic and methodological gap leaves subtropical influenza preparedness without the region-specific evidence required for precision-oriented public health planning.

A key reason subtropical weather-influenza relationships remain difficult to characterize is their non-linear, threshold-dependent, and lagged nature. Conventional linear analyses, such as logistic regression, assume simple contemporaneous relationships and thus fail to capture these dynamics with the precision required for epidemiological inference (Ng et al. 2022). Moreover, meteorological effects on influenza transmission also unfold over days to weeks, further complicating association analysis (Ng et al. 2022; Si et al. 2024). Distributed Lag Non-linear Models (DLNM) provide a framework for jointly analyzing exposure-response and lag-response relationships (Gasparrini, Armstrong, and Kenward 2010; Gasparrini 2011). DLNM has been applied in empirical analyses of influenza transmission patterns and in lagged risk analyses between influenza and meteorological factors (Liu et al. 2020; Guo et al. 2019). However, DLNM is fundamentally an inferential tool designed for retrospective association characterization and does not generate operational forecasts capable of supporting prospective public health decision-making.

On the forecasting side, traditional statistical models, such as Autoregressive Integrated Moving Average (ARIMA) approaches, despite their widespread use for influenza prediction (Wang et al. 2017), struggle to capture non-linear, lagged interactions among multiple meteorological drivers (Shoji, Katayama, and Sano 2011). Existing studies often overlook spatiotemporal heterogeneity intrinsic to subtropical contexts (Zhang et al. 2022). Although time series analysis approaches and deep neural learning architectures, such as Recurrent Neural Networks (RNNs) (Venna et al. 2018), can accommodate non-linear temporal patterns (Venna et al. 2018), their performance is hampered when baseline algorithms are not deeply customized, due to gradient vanishing or explosion and memory decay over longer horizons (Liu et al. 2021). Long Short-Term Memory (LSTM) algorithms, an advanced RNN variant, address these issues through gating mechanisms that enable learning of long-term temporal dependencies (Absar et al. 2022; Liu et al. 2021), and have shown potential in influenza forecasting (Absar et al. 2022), although prior applications have often struggled to architectural complexity, high computational cost, and typically lower interpretability than regression-based methods (Liu et al. 2021). For instance, LSTM algorithms have demonstrated good predictive performance in the Chinese subtropical Fujian region but remained limited to identify subtype-specific virological differences (Zhu et al. 2022). Another study proposed a complex seasonal autoregressive integrated moving average-LSTM (SARIMA-LSTM) hybrid utilizing singular spectrum analysis to predict influenza in Shanxi. However, its analytical complexity and computational intensity limited the explicit incorporation of confounding behavioral variables (Zhao et al. 2023).

Reviewing these literatures, three substantive gaps persist at the intersection of meteorological characterization and forecasting for subtropical influenza. First, most existing studies apply DLNM or LSTM in isolation, foregoing the potential synergy between interpretable association modeling and flexible non-linear forecasting. Second, forecasting models trained on data spanning the Corona virus disease 2019 (COVID-19) period rarely account explicitly for the epidemiological non-stationarity introduced by non-pharmaceutical interventions (NPIs) and fluctuating testing intensity, which can distort case-based signals in subtype-specific ways. Third, the choice of modeling endpoint, typically raw case counts rather than positivity, rendering them highly vulnerable to surveillance intensity biases, as counts confound underlying transmission with testing behavior, particularly during the pandemic and post-pandemic period.

To address these gaps, we developed a dual-stage analytical framework leveraging six years (2018-2023) of multi-site influenza surveillance data and detailed meteorological records from Putian, a representative subtropical coastal city in southeastern China that exemplifies the under-investigated non-megacity urban context described above. The 2018–2023 window deliberately spans a structural epidemiological transition: two pre-pandemic baseline years (2018–2019), three NPI-modulated years (2020–2022), and one post-suppression rebound year (2023). In the first stage, DLNMs characterize subtype-specific, non-linear, and lag-distributed associations between meteorological variables and influenza A and B positivity. In the second stage, a Bayesian-optimized LSTM network integrating meteorological, autoregressive, and socio-behavioral covariates is used for short-horizon forecasting, benchmarked against two covariate-matched baselines representing distinct methodological paradigms, a multivariate ARIMA model (canonical linear statistical reference) and an XGBoost gradient-boosting ensemble (leading non-deep-learning machine-learning framework), and evaluated in an independent subtropical city (Sanming) as a preliminary test of model portability. Influenza positivity rate is used as the primary modeling endpoint to mitigate testing-related surveillance bias. Our aim is to provide an interpretable, climate-informed forecasting approach for subtropical influenza that can serve as a methodological foundation for future operational surveillance developments.

## Methods

### Ethics statement

This study was institutionally approved by the Research Ethics Committee of the Putian Centre for Disease Control and Prevention (Putian CDC, approval number: Ethics Review 2020-003), Putian, and the Medicine and Biological Engineering Technology Research Centre of the Ministry of Health (MBETRC), Guangzhou, China. All procedures complied with the later amendments of the 1964 Declaration of Helsinki and relevant ethical guidelines for biomedical research. A major methodological strength of this study lies in its robust, uninterrupted longitudinal data collection framework spanning January 1, 2018, to December 31, 2023. While the analytical timeline encompasses a “retrospective” phase (January 1, 2018 – October 13, 2020) prior to formal ethical approval, and a “prospective” phase thereafter, we emphasize that this distinction represents a purely administrative demarcation regarding the timing of ethical approval. It does not reflect any shift in demographic cohorts, sentinel hospital locations, or data collection methodologies. Importantly, the historical data (2018–2020) were not subjected to the recall biases or misclassification risks typical of traditional retrospective chart reviews. Rather, they were systematically extracted from a continuously operating, highly standardized public health sentinel surveillance network. From the inception of data collection through the end of 2023, the local CDC maintained absolute uniformity in clinical influenza-like illness (ILI) definitions, nasopharyngeal swabbing procedures, and real-time reverse transcription polymerase chain reaction (RT-PCR) diagnostic assays. Consequently, the pre-2020 data possess the high-fidelity characteristics of a strict prospective cohort, ensuring unparalleled longitudinal consistency and mitigating temporal measurement bias across the entire pre-pandemic, pandemic, and post-restriction timeline.

Rigorous procedures were employed to ensure patient confidentiality and data privacy. Influenza surveillance data were fully anonymized prior to being accessed by the research team. The dataset included only essential epidemiological information (such as onset date, gender, age, and residential district) necessary for the analysis, excluding any personal identifiable information. Additionally, individual-level data on vaccination status or detailed socioeconomic profiles were not routinely collected. Any potential linkage between coded data and original patient identifiers maintained by Putian CDC was strictly inaccessible to the researchers. Consequently, the Research Ethics Committee waived the requirement for individual informed consent. Throughout the data collection process, the diagnosis of all ILI cases, information and specimen collection, and influenza virus nucleic acid testing strictly followed prevailing national standards, including the Diagnostic Criteria for Influenza (source: National Health Commission of the People’s Republic of China) and the National Influenza Surveillance Technical Specifications (source: China CDC). All participating medical professionals, CDC personnel, and laboratory technicians underwent professional training to ensure research quality. Meteorological and COVID-19 pandemic-related covariate data were sourced from publicly available databases, ensuring no privacy concerns. The anonymized data were securely managed, used solely for the stated research purposes, and were not disclosed to any third party.

### Study area

Putian city, on China’s southeast coast (latitudes 24°59’N–25°46’N; longitudes 118°27’E–119°39’E), is an exemplary site for examining the impact of meteorological factors on influenza transmission in subtropical regions. It experiences a subtropical monsoon climate with warm, humid summers and mild, dry winters. The annual mean temperature ranges 18–21°C, with summer highs up to 38°C and winter lows at 0.4°C. Annual precipitation averages 1538.8 mm, with 72% occurring from April to September. Covering 4200 km² and comprising five counties/districts with a population exceeding 3.2 million, Putian is a commercial/manufacturing hub with high population density and robust transport infrastructure, promoting mobility and social interactions that drive influenza transmission. Since 2004, Putian CDC has operated a comprehensive influenza surveillance network, meeting national standards, employing standardized RT-PCR testing for influenza A/B since late 2017, supported by a trained workforce, ensuring high-quality longitudinal data for robust analyses. Together, Putian’s climatic variability, socio-economic characteristics, and surveillance capacity position it as a representative site for this study, with insights potentially generalizable to other subtropical settings with similar environmental and epidemiological traits. Sanming city, between latitudes 25°30’N and 27°97’N and longitudes 116°22’E and 118°39’E, is a mountainous subtropical area about 330 km from Putian and was selected as an external validation data source.

### Study design

This collaborative study by Putian CDC and MBETRC utilized a prospective cohort design, with retrospective analysis of historical data to systematically assess non-linear quantitative relationships between multiple meteorological factors and both influenza A and B infection risks, ultimately constructing a predictive network for outbreaks. The technical workflow is illustrated in Figure 1.

**Figure 1.**
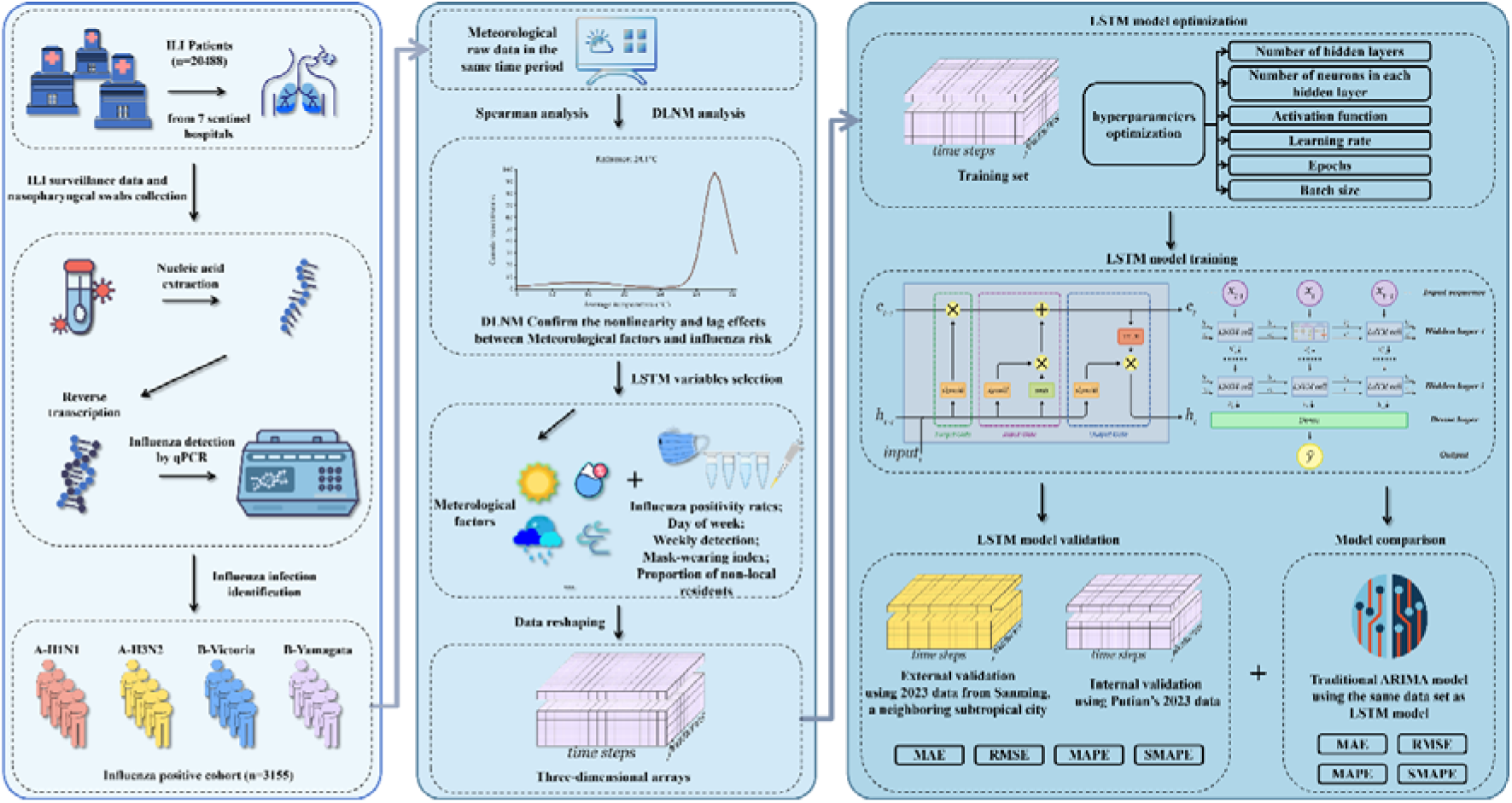
Schematic of the integrated analytical framework. Sentinel surveillance data, daily meteorological records, and contextual covariates (weekly testing volume, weekday/weekend status, proportion of non-local residents, mask-wearing stringency indices) were jointly analysed using two complementary approaches: (i) distributed lag non-linear models (DLNM) to characterise non-linear, lagged exposure–response relationships between meteorological variables and subtype-specific influenza positivity, providing mechanistic interpretability; and (ii) a Bayesian-optimized Long Short-Term Memory (LSTM) neural network for short-term forecasting of daily positivity rates, benchmarked against a seasonal ARIMA model. The primary outcome variable for both analytical streams is the influenza positivity rate (laboratory-confirmed cases ÷ ILI specimens tested), selected to mitigate surveillance bias arising from temporal variation in testing intensity.

The study spanned January 1, 2018, to December 31, 2023, integrating four categories of data sources: (i) ILI and laboratory-confirmed cases from seven influenza sentinel hospitals across Putian’s urban and rural areas, ensuring representative coverage of diverse healthcare-seeking populations; (ii) daily meteorological data; (iii) COVID-19 associated public health intervention indicators (mask-wearing stringency indices) recorded from January 2020 onwards; and (iv) demographic mobility indicators (non-local population proportion) obtained from ILI consultation records. To ensure consistency between meteorological measurements and influenza incidence records across all data sources, we applied rigorous quality control and pre-processing procedures, including: temporal alignment of all data streams to a unified daily resolution; missing value imputation using temporally adjacent observations for sporadic gaps (<5% of records); cross-validation of laboratory-confirmed cases against ILI consultation records to identify and resolve coding inconsistencies; and standardization of meteorological measurements against the regional monitoring network’s established calibration protocols. Final datasets underwent independent verification by two co-investigators to ensure analytical reliability.

DLNM was first constructed to screen meteorological factors and other covariates with substantial influence on influenza seasonality. Subsequently, an LSTM neural network was developed within the same covariate system. The integrated DLNM–LSTM framework was employed as methodologically complementary rather than redundant components, leveraging the distinctive strengths of each approach to address different analytical objectives within a unified investigation. Specifically, DLNM models characterize the non-linear exposure–lag–response relationships between meteorological factors and influenza risk, providing biologically interpretable insights into the temporal structure of weather-influenza associations and identifying meteorological factors with statistically and clinically significant effects on influenza dynamics. Building upon this DLNM-derived foundation, the LSTM network constructs a time-series forecasting tool within an identical covariate framework, evaluating predictive capability for influenza transmission trends. Beyond meteorological factors, our DLNM-LSTM framework systematically incorporated influenza positivity rates as the primary outcome variable, weekly detection volumes, mask-wearing stringency indices (indicator of NPIs during the COVID-19 pandemic), day of the week (DOW, distinguishing weekdays from weekends), and non-local population proportion (defined as the ratio of the number of individuals whose reported residential district at the time of testing lies outside Putian city to the total number of tests) as covariates within both DLNM and LSTM modeling pipelines. This comprehensive covariate framework ensures that observed meteorological associations are estimated after controlling for surveillance intensity, public health intervention status, behavioral healthcare-seeking cycles, and population mobility patterns. The LSTM network was trained on data from Putian corresponding to the four categories described above, with the time period 2018-2022, and Putian’s 2023 data serving as the internal validation set. To rigorously evaluate model transferability beyond the training context, data from Sanming city, a mountainous subtropical city exhibiting comparable climatic characteristics, influenza seasonality, and public health intervention frameworks to Putian, were acquired for the period January 1, 2023, to December 31, 2023, temporally aligned with the Putian internal validation set. This Sanming dataset constituted our external validation set, facilitating a geographically independent assessment of model generalization within subtropical Chinese environments. The predictive performance of the LSTM algorithm for influenza A/B prevalence in 2023 Putian data was benchmarked against two parallel baseline models: (1) the multivariate ARIMA model with exogenous variables (ARIMAX), representing the canonical linear statistical reference in public-health surveillance; and (2) the XGBoost gradient-boosting regression model, representing the leading non-deep-learning machine-learning framework for structured predictive tasks. To ensure paradigm-agnostic methodological comparisons, both baseline models were supplied with the exact same meteorological and epidemiological covariate matrix as the LSTM, without any additional hand-engineered lag features, sliding-window statistics, or autoregressive terms in the XGBoost input.

### Study cohort

The study systematically screened outpatients and emergency departments across seven influenza surveillance sentinel hospitals within Putian’s prefecture, with an emphasis on high-incidence departments such as fever clinics, internal medicine, and pediatrics. The screening encompassed ILI cases from January 1, 2018, to December 31, 2023. According to earlier mentioned Diagnostic Criteria for Influenza, ILI cases are defined as acute onset with high fever (≥38°C) accompanied by respiratory symptoms such as cough or sore throat yet lacking a definitive causative diagnosis. Respiratory samples, together with onset dates, ages, genders, and residence information, were collected from all ILI patients. Unified diagnostic standards, laboratory testing methods, and quality control measures were employed throughout the research process to ensure the reliability and comparability of results.

After conducting nucleic acid screening for influenza viruses from the collected respiratory samples of ILI cases, the criteria for inclusion in this study prescribed the following: 1) the patient presented ILI symptoms; 2) the nucleic acid test for the influenza virus yielded a positive result; 3) the patient is a permanent resident of Putian city; 4) the duration of symptoms from onset to consultation did not exceed three days. No restrictions were placed on age or gender. The exclusion criteria were: 1) patients with clear alternative diagnoses, such as bacterial pneumonia or tuberculosis; 2) patients with unstable underlying conditions, such as acute exacerbations of chronic obstructive pulmonary disease (COPD); 3) patients with missing data. This focused the analysis on acute ILI presentations.

### Sample collection and pathogen typing

Respiratory specimens (nasopharyngeal swabs) were collected from ILI patients during their initial visit, prior to treatment, and stored at 4°C in viral transport medium. All samples were delivered to the Putian CDC laboratory within 24 hours of collection. Nucleic acid extraction and one-step real-time fluorescent RT-PCR for influenza A/B subtyping (H1N1, H3N2, Victoria, and Yamagata lineages) were executed within 24 hours upon sample arrival. All assays utilized commercial diagnostic kits (supplied by Da’an Gene Co., Ltd., Guangzhou, China) and were processed in a Biosafety Level 2 (BSL-2) laboratory, strictly adhering to the manufacturer’s instructions and China CDC’s standardized protocols. Detailed laboratory procedures, including RNA integrity parameters and PCR amplification thresholds, are comprehensively documented in the Supplementary Information (Additional file 1).

### Meteorological and mask-wearing stringency data sources

Daily meteorological observation data were sourced from the authoritative public database Visual Crossing (https://www.visualcrossing.com/), which provides extensive historical time-series data across multiple meteorological variables on a global scale. We extracted meteorological variables for Putian city, including daily average temperature (°C), relative humidity (%), precipitation (mm), diurnal temperature range (°C), wind speed (km/h), wind direction, solar radiation (W/m^2^), and the ultraviolet (UV) index, wherein the diurnal temperature range was defined as the difference between the highest and lowest temperatures of the day. All indicators were synchronized with the influenza data. To ensure data quality, we conducted missing value and anomaly checks on the raw datasets.

Early COVID-19 studies revealed a notable decline in positivity rates of seasonal respiratory viruses relative to pre-pandemic levels, likely due to containment measures such as masking and behavioral changes during surges (Groves et al. 2021; Xiao et al. 2022). To evaluate the potential impact of NPIs on influenza incidence, we obtained policies on face-covering stringency indices for China during the same period from the Our World in Data (OWID) database (https://ourworldindata.org/coronavirus) as a reference indicator for social prevention measures. This database categorizes stringency indices into four levels: 0 represents “No policy”; 1 represents “Recommended”; 2 represents “Required in some specified shared/public spaces outside the home with other people present, or some situations when social distancing not possible”; 3 represents “Required in all shared/public spaces outside the home with other people present or all situations when social distancing not possible”; 4 represents “Required outside the home at all times regardless of location or presence of other people”. While OWID stringency indices represent national-level policy, publicly accessible and standardized daily NPI datasets at the municipal level are currently unavailable in China. To ensure the spatial validity of these indices, co-authors from Putian CDC, who actively managed the local epidemic response, conducted a rigorous cross-validation. Our local public health experts confirmed that temporal fluctuations of OWID indices (ranging from Level 4 strict mandates in 2020 to Level 1 recommendations in 2023) exhibited high fidelity with the actual, on-the-ground enforcement of NPIs in both Putian and Sanming. Thus, OWID stringency indices serve as a highly reliable contextual proxy for local social contact restrictions. The datasets from these public databases, along with influenza surveillance data, are devoid of missing values, eliminating the need for imputation. For influenza and COVID-19 data, days with no reported cases were explicitly recorded as zero per the sentinel hospitals’ and government’s daily reporting protocols, respectively.

### DLNM analysis

In the DLNM stage, we prioritized epidemiological interpretability by focusing on the isolated, non-linear, and lag-distributed main effects of individual meteorological variables. We deliberately avoided incorporating high-dimensional interaction cross-bases (e.g., temperature × humidity interactions) within the DLNM to prevent the “curse of dimensionality,” which typically induces severe multicollinearity, variance inflation, and loss of interpretability. Instead, the computational task of capturing complex, synergistic interactions among concurrent meteorological factors and epidemiological covariates was entirely delegated to the subsequent LSTM network. Separate DLNM models were constructed for influenza A and B incidence risks. To rigorously address the inherent confounding effects of “surveillance intensity bias”, where fluctuations in raw case counts may merely reflect transient surges in clinical testing capacity rather than true community transmission, the primary outcome metric for all DLNM modeling was mathematically defined as influenza positive rates, with meteorological factors serving and weekly detection volumes as independent variables. The formula for calculating the daily influenza positivity rate is as follows:

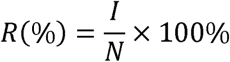

where *R* represents the daily influenza positivity rate, *I* is the number of daily influenza positive cases, and *N* denotes the total number of daily influenza tests performed. By adopting this WHO-recommended surveillance metric, our analytical framework explicitly standardizes the epidemiological denominator. This mathematical normalization effectively neutralizes the severe surveillance intensity bias caused by dramatic testing volume surges during the COVID-19 pandemic, ensuring that our models capture intrinsic viral transmissibility rather than artificial fluctuations in healthcare-seeking behavior or diagnostic capacity.

The maximum lag time was set to 15 days for all models, optimizing biological validity and analytical precision to capture typical influenza incubation and transmission cycles while minimizing model complexity and statistical noise from extended periods, aligning with previous research. A quasi-Poisson function was used to account for overdispersion in the incidence data. The model formula is as follows:

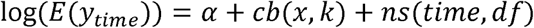

Where α is the intercept, *cb* is the cross-basis function of the meteorological factor *x*, which includes the exposure-response relationship ns(x, df_x_) defined using natural splines and the lag-response relationship g_(k)_ defined using polynomial functions, with *k* as the lag time; ns(time,df_time_) is a natural spline function reflecting long-term trends, used to control for long-term trends and seasonality. The degrees of freedom *df_x_* for the exposure-response relationship nsspline were selected from the range {2, 3, 4, 5}, while the degrees of freedom *df_time_* per year for the long-term trend ns spline were selected from the range {6, 7, 8, 9, 10}. To determine the optimal *df* , including *df_x_* for the natural spline modeling the exposure-response relationship and *df_time_* per year for the natural spline modelling the long-term trend, the combinations of above ranges across selected meteorological factors were systematically evaluated using the quasi-Akaike information criterion (QAIC), with the combination yielding the smallest QAIC being selected. This indicates the best balance between goodness-of-fit and model complexity for our influenza-specific dataset. The QAIC formula is:

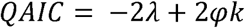

where *λ* represents the log-likelihood of the model, *φ* is the dispersion parameter, and *k* indicates the number of model parameters. When plotting prediction results, the median was selected as the reference value for its robustness against outliers, offering a consistent depiction of typical meteorological conditions in Putian and enabling the evaluation of bidirectional risk deviations, especially during extreme weather events. To further explore the impact of extreme weather conditions, we predicted and displayed the effects of extreme scenarios across the meteorological factors: except for precipitation extremes set at 5 mm and 25 mm and wind direction extremes at the most common eastern and rarest northwestern winds, all other meteorological factors were assessed at their 5^th^ and 95^th^ percentiles to estimate the lagged risk curves relative to the median. All results underwent seasonality and long-term trend corrections to eliminate the inherent temporal variation effects between meteorological factors and influenza incidence. Furthermore, to evaluate the robustness of our findings against testing intensity, we conducted a targeted sensitivity analysis by reconstructing the DLNM models without the “weekly detection volumes” covariate. We then compared the non-linear cumulative risks and extreme weather lag effects between these ablated models and the original fully adjusted models. This allowed us to determine whether the identified climate-transmission associations were stable and biologically intrinsic, or merely artifacts of weather-correlated testing behaviors (Supplementary Information Additional file 4).

### LSTM modeling and analysis

Building on the significant non-linear and lagged effects through DLNM analysis of real-world environmental exposures, this study further constructed multi-factor influenza A and B prediction LSTM networks. Leveraging a recurrent architecture with non-linear gating mechanisms, these networks are able to automatically capture and represent complex, high-dimensional interactions among meteorological variables without the need for manual pre-specification. This functional division of labor between the DLNM and LSTM models enhances predictive performance while preserving the interpretability and inferential stability established in the DLNM stage.

### Data preprocessing

To respect the temporal dependence inherent in LSTM architectures and avoid data leakage, we adopted a strict chronological out-of-time (OOT) validation strategy. Time-series data from January 1, 2018, to December 31, 2022 (88.26% of the Putian dataset) were used for model training, with 10% reserved during Bayesian optimization as an internal validation subset for convergence monitoring and hyperparameter tuning only. Data from January 1, 2023, to December 31, 2023 (11.74%) were held out as a chronologically internal validation set for final performance evaluation, covering a complete annual cycle. External validation was conducted using concurrent 2023 data from Sanming city to assess spatial generalizability. To prevent distributional leakage, all normalization parameters were derived exclusively from the training set and consistently applied to the validation sets, with predictions subsequently transformed back to the original scale. First, we performed data normalization, a crucial step to ensure that training and test set data are compared on a unified scale. We normalized the training and test set data separately within the range [0, 1]. For the test set normalization, we used the maximum and minimum values from the training set as boundaries. This approach ensured consistency between the normalized test set data and the training set data. After the algorithm conducted predictions on the test set data, we performed denormalization to convert the predicted results back to the original data scale and rounded them to integers. These steps ensured that the final prediction results accurately and objectively reflected the LSTM’s performance in real-world scenarios and provided reliable data for subsequent calculation of evaluation metrics.

The normalization formula is as follows:

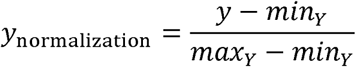

The denormalization formula is as follows:

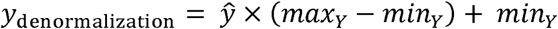

where *y_normalization_* is the normalized result, *y* is the original data, *max_Y_* is the maximum value of the variable in the training set, *max_Y_* is the minimum value of the variable in the training set, *̂y* is the predicted value output by the network, and *y*_denormalization_ is the denormalized result.

Finally, we used the above data while account for potential confounders by incorporating features such as DOW (1 for Monday–Friday, 0 for Saturday–Sunday), weekly detection volumes, non-local population proportion, and mask-wearing stringency indices, as input variables. The time step was set to 15 (i.e., using data from the previous 15 days to predict the number of influenza cases for the following day), and the two-dimensional data were organized into three-dimensional arrays to serve as input for the LSTM network.

### Network construction

Our algorithm is a unidirectional LSTM, using Adam as the optimizer and Mean Square Error (MSE) as the loss function. The number of hidden layers was determined through hyperparameter optimization to ensure optimal network performance. Each LSTM layer consists of multiple LSTM units, including input gates, output gates, and forget gates. The basic framework of LSTM and the structure of LSTM units are shown in the LSTM model training section of Figure 1. Detailed principles and formulas are provided in the LSTM cell description section of Supplementary information and illustrated in Figure S1.

In the LSTM network, we used Hyperopt for hyperparameter tuning, with MSE established as the reference metric for evaluating and selecting optimal parameters. A detailed description of the Hyperopt Bayesian optimization strategy, including the specific search spaces for learning rates, network architecture (layers and neurons), and activation functions, along with their associated theoretical justifications, is provided in Supplementary Information (Additional file 1).

### LSTM performance evaluation

To comprehensively assess the predictive performance of the LSTM network, this study employed multiple commonly used error metrics, including Mean Absolute Error (MAE), Root Mean Square Error (RMSE), Mean Absolute Percentage Error (MAPE), and Symmetric Mean Absolute Percentage Error (SMAPE) (Chicco, Warrens, and Jurman 2021).

MAE reflects the actual magnitude of the error between predicted and true values. A smaller MAE indicates more accurate network predictions. The calculation formula is:

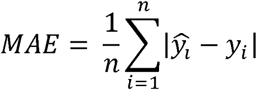

RMSE is the arithmetic square root of the mean of the sum of squared errors. A smaller RMSE indicates better network prediction performance. The calculation formula is:

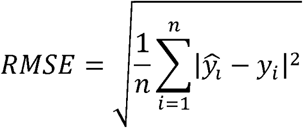

A smaller MAPE indicates better network fitting. Since the denominator cannot be zero, this study only calculated MAPE for data where the true value was not zero. The calculation formula is:

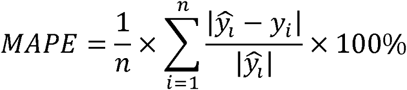

A smaller SMAPE indicates better network fitting. The calculation formula is:

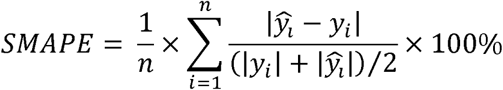

In the above formulas, *̂y_l_* is the denormalized predicted value, *y_i_* is the observed value, and *n* is the number of observations.

Additionally, we plotted the loss function curves for the network on the training and validation sets to monitor LSTM convergence and the risk of overfitting. By comparing the trends and differences between the two curves, we could intuitively judge the training quality and generalization ability of the LSTM. Furthermore, the Bayesian-optimized LSTM architecture was directly applied, without re-training or hyperparameter adjustment, to both the Putian internal test set and the Sanming external validation set. This unified modeling framework ensures that performance differences between the two evaluation contexts primarily reflect geographic transferability rather than model re-optimization.

To further explain the prediction logic of the LSTM, this study employed the SHapley Additive exPlanations (SHAP) method for interpretability analysis. Based on Shapley values from game theory, SHAP can quantify the contribution of each feature to the LSTM’s predicted values and generate intuitive feature importance rankings and dependency plots. Positive values indicate that the feature increased the predicted result, while negative values indicate a decrease in the predicted result, with the absolute value representing the intensity of the influence. SHAP values were computed by treating each meteorological variable’s lagged sequence as a unified input feature, preserving the temporal dependencies within each sequence rather than decomposing lags into independent features. This approach not only aligns with the sequential nature of our LSTM inputs but also supports our focus on population-level trends rather than individual-level causal effects.

To compare the predictive performance between the LSTM and alternative forecasting paradigms, we concurrently developed two baseline models, the ARIMA model and the XGBoost machine-learning model, employing the same training and validation datasets. Detailed methodological description for ARIMA and XGBoost are provided in Supplementary Information Additional files 5 and 6, respectively. To assess the contribution of pandemic-related confounding variables, we performed targeted sensitivity analyses on the LSTM networks. Specifically, we sequentially removed covariates of mask-wearing stringency indices and weekly detection volumes from the input features while maintaining identical Bayesian-optimized hyperparameters. The performance of these ablated models was evaluated using MAE, RMSE, MAPE, and SMAPE, allowing us to quantify the exact necessity of incorporating testing and behavioral covariates in forecasting models during periods of epidemiological non-stationarity.

### Statistical analysis

Statistical analyses were primarily conducted using R software version 4.3.1 and Python version 3.8.3. DLNM modeling was implemented using the DLNM package in R. LSTM construction used Python 3.8.3, with the main program packages including tensorflow 2.2.0, keras 2.3.1, and shap 0.44.1. The ARIMA model was developed using the forecast package in R. The XGBoost model was implemented using the xgboost package (version 3.2.1.1) in R, with hyperparameter tuning conducted via grid search over tree depth, learning rate, and L2 regularization parameters; detailed specifications are provided in Additional file 6. All statistical tests were two-sided, with P < 0.05 considered statistically significant.

## Results

### Descriptive epidemiological characteristics of influenza

Between 2018 and 2023, influenza surveillance conducted at seven sentinel hospitals in Putian city documented a total of 20,488 ILI cases, with the majority being influenza-negative (17,333 cases, 84.60%). Of these, 3,155 cases were confirmed positive for influenza through PCR testing, representing a positivity rate of 15.40%. By viral type, 1,685 cases were attributed to influenza A (8.23%), and 1,470 cases were influenza B (7.17%). Further subtype analysis of influenza A revealed 790 cases of H1N1 (46.88%) and 895 cases of H3N2 (53.12%), while for influenza B, 1,270 cases were of the Victoria lineage (86.39%) and 200 cases were of the Yamagata lineage (13.61%). Notably, following the onset of the COVID-19 pandemic in the spring of 2020, there was a significant reduction in the incidence of influenza. The annual incidence rates for influenza in 2020 and 2021 were 4.54 per 100,000 and 5.59 per 100,000, respectively, markedly lower than the pre-pandemic rates observed in 2018 (16.01 per 100,000) and 2019 (18.74 per 100,000), suggesting that public health measures implemented during the COVID-19 pandemic, such as mask-wearing and social distancing, were highly effective in decreasing the transmission of respiratory infections, including influenza.

In terms of seasonal distribution, surveillance data (Figure 2 A) revealed a notable dynamic interplay between influenza A and B viruses over the study period. In 2018, influenza A H1N1 was the dominant strain. However, by 2019, cases of the influenza B Victoria lineage gradually increased, surpassing H1N1 during the summer to become the predominant strain. In 2020, there was a sharp decline in influenza cases, with influenza A H3N2 emerging as the leading strain. The dominance of influenza B Victoria returned in 2021, only to be overtaken by a resurgence of influenza A H3N2 in the summer of 2022. By the winter of 2023, both influenza A H3N2 and influenza B Victoria exhibited concurrent surges, highlighting a dual epidemic. Interestingly, contrary to the conventional understanding that influenza A typically drives larger epidemics, our findings show that influenza B accounted for 52.18%, 100%, and 64.76% of total influenza cases in 2019, 2021, and 2022, respectively.

**Figure 2.**
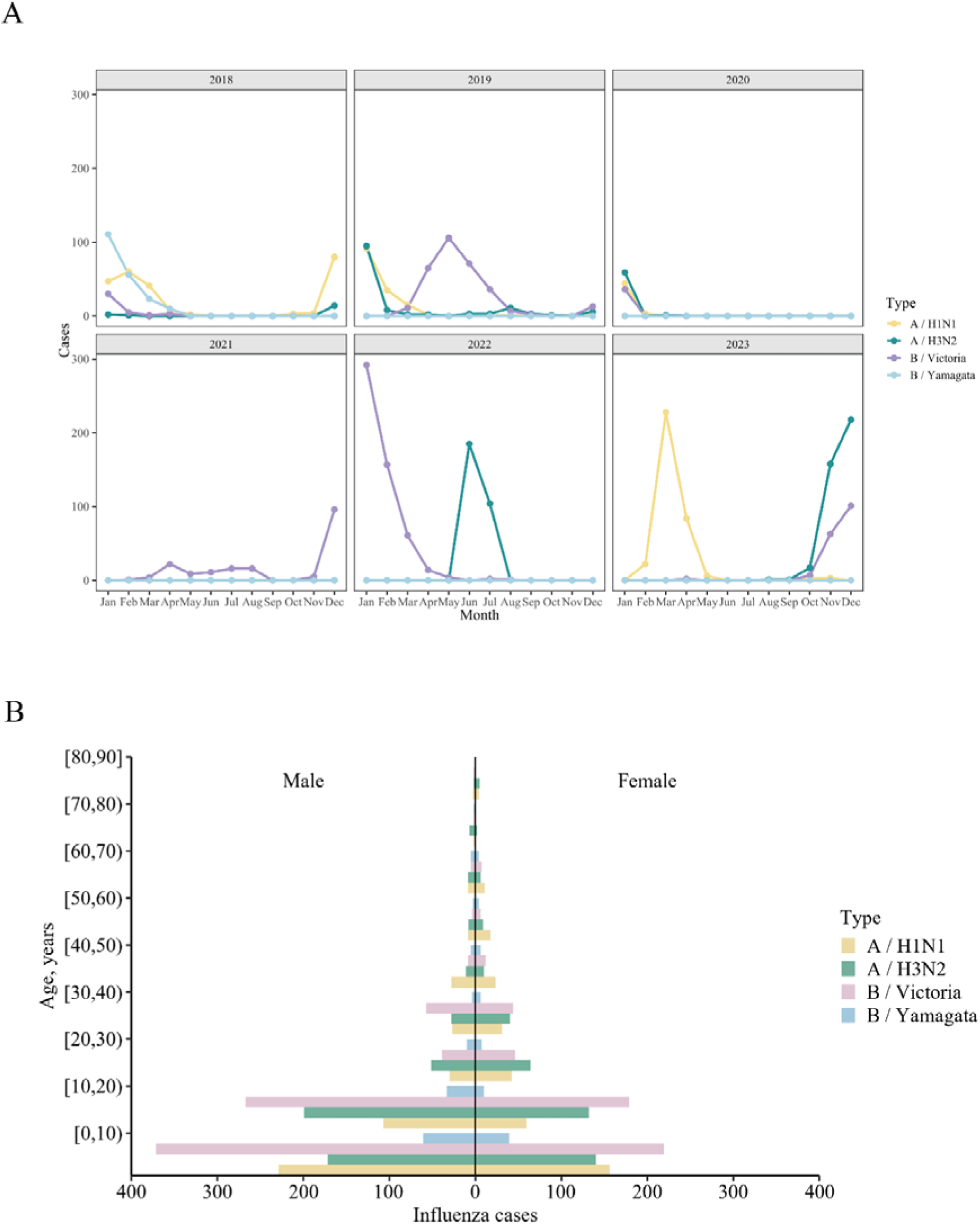
Temporal trends and demographic distribution of confirmed influenza cases in Putian city (2018 to 2023). This figure presents the temporal and demographic characteristics of influenza cases in Putian City over the period from 2018 to 2023. **(A)** Line plots illustrating the monthly case counts of various influenza subtypes (Influenza A: H1N1, H3N2; Influenza B: Victoria, Yamagata) across the six-year study period. The transition from historical heatmaps to precise line plots enhances the visualization of distinct epidemiological surges, notably capturing the profound interruption of typical seasonal transmission during the peak COVID-19 pandemic restrictions (2020–2021) and the subsequent viral rebound. **(B)** A population pyramid detailing the age and sex distribution of the 3,155 laboratory-confirmed influenza cases. The visualization bifurcates cases by gender (males on the left, females on the right) across age intervals, underscoring the differential susceptibility of specific demographic cohorts, particularly highlighting the heightened vulnerability of children (0-10 years) and the elderly (60 years and above).

In the demographic analysis (Figure 2 B), among the 3155 laboratory-confirmed influenza cases, males (1,811 cases; 57.40%) outnumbered females (1,344 cases; 42.60%), with a male-to-female ratio of 1.35:1. Significant gender differences were observed across age groups (χ² = 37.36, P < 0.001), with males constituting 60.68% of cases in individuals under 18 years, whereas females were more prevalent among adults (51.53%). A statistically significant difference was noted in the gender distribution between influenza A and B (χ² = 6.64, P = 0.01), with males comprising a higher proportion (59.86%) of influenza B cases. No statistically significant gender differences were found in the distribution of influenza subtypes (χ² = 2.43, P = 0.49). Additionally, children and adolescents under 18 years of age were identified as the most at-risk population, comprising 73.12% of all cases. Influenza B was more prevalent in this age group (50.33% vs. 49.67%), while influenza A predominated in the adult population (63.56% vs. 36.44%). These findings suggest that susceptibility to different influenza virus types may vary across age groups.

Overall, our findings suggest that in subtropical regions, the epidemic intensity of Influenza B may rival or even exceed that of Influenza A. Furthermore, the widespread implementation of non-pharmaceutical interventions (e.g., mask-wearing) during the COVID-19 pandemic significantly curtailed the transmission of respiratory viruses, including influenza. Continuous monitoring of Influenza A (H1N1, H3N2) and Influenza B Victoria strains is essential, alongside heightened surveillance and preventive measures targeting children and adolescents, particularly in densely populated settings such as schools. These insights are crucial for informing vaccine strain selection, vaccination strategies, and targeted public health interventions.

### Temporal trends in meteorological factors and influenza incidence

Between 2018 and 2023, influenza incidence in Putian city exhibited distinct seasonal patterns. The majority of cases occurred between December and April of the following year, accounting for over 60% of the annual cases, with a secondary peak observed from May to September. Concurrently, meteorological factors demonstrated clear cyclical variations (Figure 3), with detailed descriptive statistics provided in Supplementary Table S1. During the summer months (July–August), the average daily temperature peaked above 32°C, while the winter months (December to February) experienced the lowest temperatures, dropping below 10°C (Figure 3 A). This period was also marked by a significant increase in diurnal temperature variation, with the maximum daily range exceeding 13°C (Figure 3 E). The annual average relative humidity was approximately 76.10%, with the highest levels observed during the monsoon season from April to June, reaching or exceeding 97% (Figure 3 B). Precipitation showed a pronounced seasonal distribution, peaking from June to September, while the period from November to February was characterized by markedly reduced rainfall (Figure 3 C). Solar radiation (Figure 3 D) and UV index (Figure 3 F) both reached their annual maxima between July and September. High wind speeds were frequently observed in Putian city from July to September during the study period, which may correlate with the increased frequency of typhoons during this period (Figure 3 G). Throughout the year, the easterly wind direction was predominant, with infections in every month (Figure 3 H).

**Figure 3.**
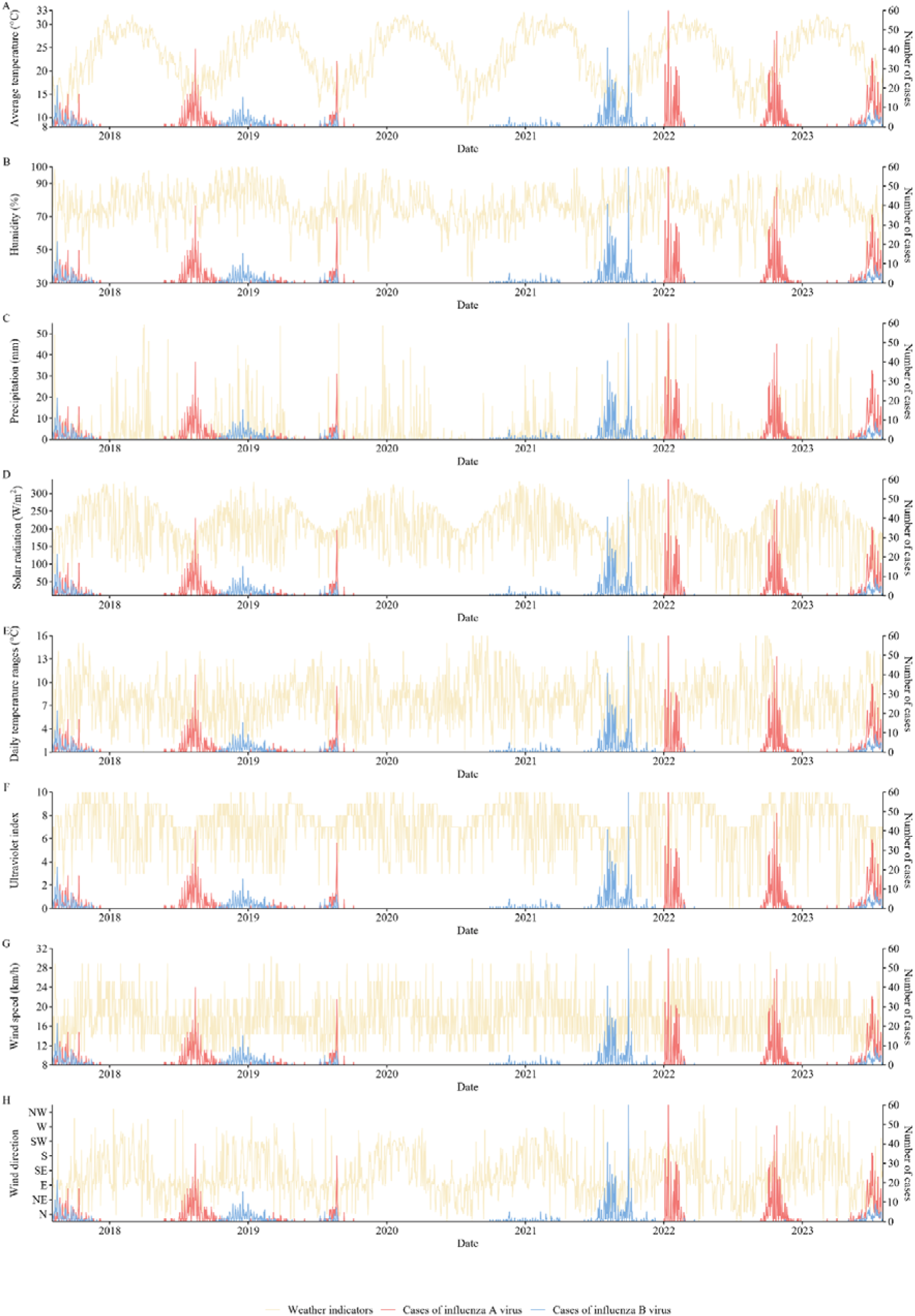
Time-series dynamics of meteorological factors and absolute influenza incidence in Putian city from 2018 to 2023. This figure vertically aligns temporal fluctuations of influenza-related and key meteorological variables, January 2018-December 2023. From top to bottom: **(A)** average temperature, **(B)** humidity, **(C)** precipitation, **(D)** solar radiation, **(E)** daily temperature ranges, **(F)** ultraviolet index, **(G)** wind speed, and **(H)** wind direction, plotted continuously over the five-year surveillance period. In each panel, the left y-axis corresponds to the observed values of the respective meteorological factor (yellow lines), while the right y-axis delineates the absolute number of confirmed cases for Influenza A (red lines) and Influenza B (blue lines). These descriptive plots visually contextualize the raw seasonal correlations and climatic extremes preceding major influenza outbreaks. Note the substantial suppression of influenza activity during 2020–2022 (pandemic intervention period) and the pronounced post-restriction rebound during 2023, particularly for influenza A, which exceeded pre-pandemic peak levels.

A comparative analysis of the temporal patterns of influenza cases and meteorological factors revealed several noteworthy correlations. Firstly, peaks in influenza A and B cases were predominantly observed during January to March, characterized by lower average temperatures and larger diurnal temperature ranges (Figures 3 A and E). Interestingly, influenza B occasionally exhibited summer outbreaks, indicating a potentially greater adaptability to higher temperatures. Secondly, extreme rainfall events and periods of high humidity were often temporally aligned with short-term surges in influenza A and B cases (Figures 3 B and C). For instance, the peak in influenza B cases in June 2022 coincided with an episode of extreme rainfall. Moreover, the majority of influenza outbreaks occurred during periods of reduced solar radiation and lower UV indices (Figures 3 D and F). Variations in wind speed were not significantly correlated with influenza incidence in our analysis (Figure 3 G). During periods of easterly winds, there was a noticeable decrease in the incidence of influenza A and B cases in Putian city (Figure 3 H). Our findings indicate that the incidence of influenza in the subtropical city of Putian was closely associated with various meteorological conditions.

### DLNM analysis for the cumulative and lagged effects of multiple meteorological factors on influenza incidence risks

Prior to analytical modeling, we assessed the pairwise associations and potential collinearity among all meteorological variables using a comprehensive correlation heatmap and scatterplot matrix (Supplementary Figure S2). Notably, certain variables, such as solar radiation and the UV index, exhibited high positive correlations. To avoid coefficient instability typically caused by multicollinearity in regression models, all DLNM analyses adopted a univariate approach for meteorological factors, sequentially evaluating the non-linear and lagged effects of individual meteorological predictors while adjusting for time trends and including other fixed covariates. Conversely, all variables were retained during the LSTM forecasting phase, as the non-linear gating architecture of recurrent neural networks inherently exhibits robust regularization against potential collinearity among input features.

To elucidate the non-linear associations between meteorological factors and influenza transmission dynamics, DLNM models were constructed using influenza positivity rates as the outcome variable. For each meteorological variable, the median exposure level was used as the reference. For each influenza-specific DLNM model, the combination of *df_x_* for the exposure-response spline and *df_time_* for the long-term trend spline that yielded the minimum QAIC was selected as the optimal configuration and applied to the final model. Detailed QAIC values for all tested combinations are provided in Supplementary Table S2. Overall, the associations between the eight evaluated meteorological factors and the cumulative risk of influenza positivity exhibited pronounced non-linear and subtype-specific characteristics over the 15-day lag period (Figure 4 A and B). Influenza A was primarily associated with lower temperature, reduced diurnal temperature range, and intermediate UV levels, whereas influenza B showed broader meteorological sensitivity, with increased risks observed under lower temperature, higher humidity, lower solar radiation, lower UV index, and specific wind directions.

**Figure 4.**
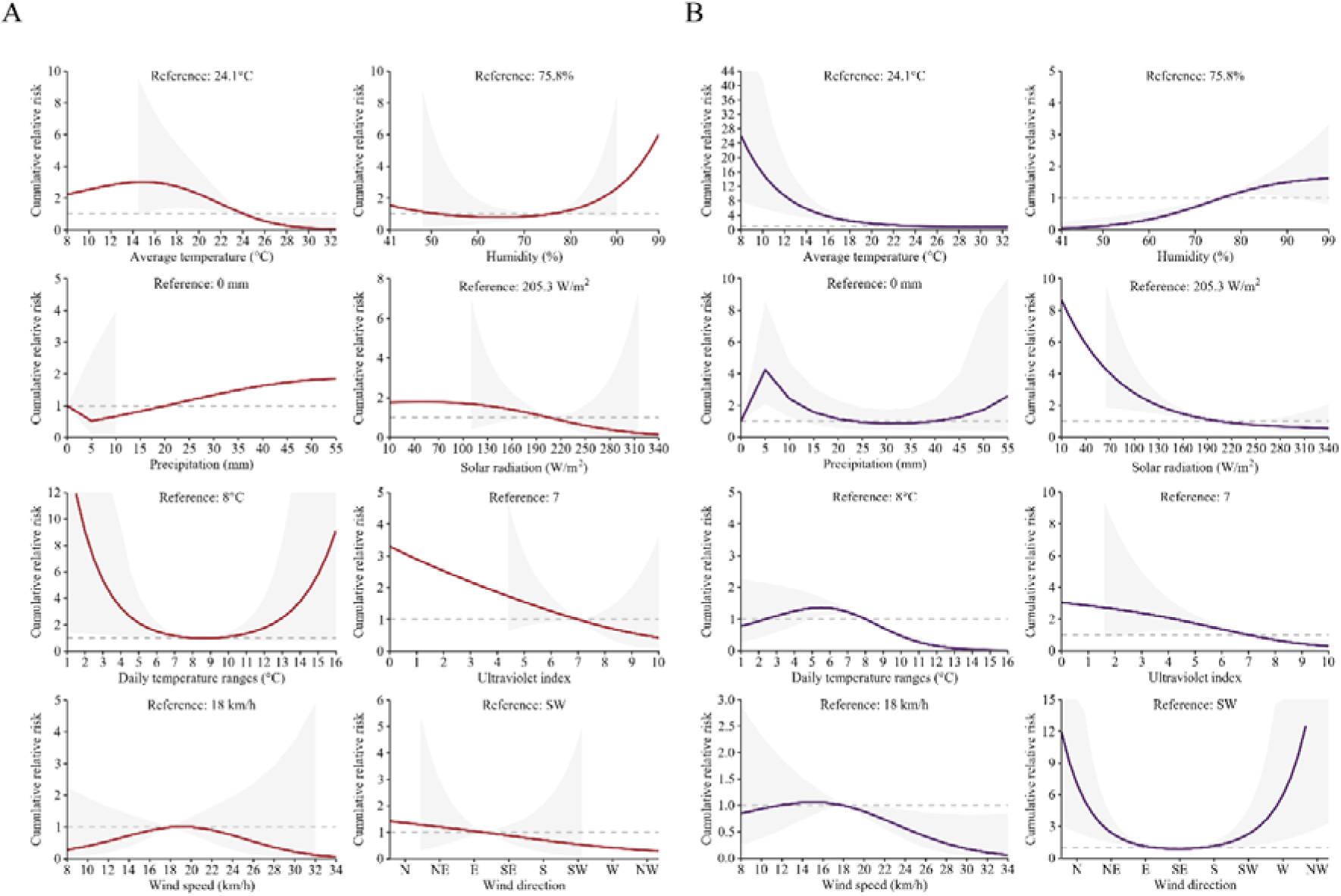
Cumulative risk of meteorological factors on Influenza A and B positivity rates over a 15-day Lag period. This figure illustrates the non-linear, cumulative exposure-response relationships between eight individual meteorological drivers and the 15-day cumulative relative risk of influenza A and B positivity, estimated via Distributed Lag Non-linear Models (DLNM). Models were adjusted for long-term trends, weekly detection volumes, and other fixed covariates. Panels in **(A)** depict the non-linear cumulative risks for Influenza A, while panels in **(B)** shows the corresponding risks for Influenza B. In each subplot, the x-axis represents the observed continuous range of a specific meteorological factor, and the y-axis indicates the cumulative relative risk compared to a predefined median reference value (e.g., 24.1°C for average temperature, 75.8% for humidity). The solid line reflects the estimated non-linear trend of risk variation, and the shaded regions denote the 95% confidence intervals (CIs).

Regarding temperature effects, using 24.1°C as the reference, the cumulative risk for influenza A showed a non-linear inverse-U pattern, peaking at 15°C with a relative risk (RR) of 3.01 (95% CI: 1.03-8.78) and gradually decreasing across the 15–24°C range. In contrast, the cumulative risk for influenza B peaked at a much lower temperature of 8°C (RR = 26.12, 95% CI: 7.79–87.61), with the risk steadily declining between 8°C and 24°C.

Humidity demonstrated divergent effects between the two subtypes. While relative humidity showed no significant association with influenza A risk (using 75.8% as a reference), the cumulative risk for influenza B increased gradually within the high-humidity range of 76% to 91%, reaching its peak at 91% (RR = 1.52, 95% CI: 1.00–2.29). Similarly, precipitation showed no significant impact on influenza A. However, for influenza B, precipitation within the 0–10 mm range exhibited a rise-then-fall risk pattern, peaking at 5 mm (RR = 4.25, 95% CI: 2.11–8.58). These findings suggest that influenza A is predominantly driven by specific temperature bands, whereas influenza B transmission is more closely linked to cool-wet meteorological regimes.

Solar radiation and the UV index also manifested distinct subtype affinities. For influenza A, neither low (75 W/m²) nor high (305 W/m²) solar radiation yielded significant cumulative risks. In contrast, low solar radiation (75 W/m²) posed a significant risk for influenza B, peaking at 9.6 days (RR = 1.16, 95% CI: 1.08–1.23). For the UV index (reference = 7), influenza A only showed significant risk exactly at an index of 7 (RR = 1.00, 95% CI: 1.00–1.00), whereas influenza B risk progressively decreased within the UV index range of 2.6 to 7, peaking at 2.6 (RR = 2.49, 95% CI: 1.04–5.96).

Diurnal temperature range (DTR) was associated with both influenza subtypes, although the estimates for influenza A were imprecise. Relative to 8°C, the cumulative risk of influenza A was highest at 1°C (RR = 15.69, 95% CI: 1.42–173.00), while influenza B risk peaked at 5.5°C (RR = 1.36, 95% CI: 1.09–1.69). Wind speed was not significantly associated with either influenza subtype. Wind direction was not associated with influenza A, but northerly, easterly, and westerly winds were associated with elevated influenza B risk, with the highest estimate observed for northerly winds (RR = 27.33, 95% CI: 3.33–224.41).

### Lagged effects of extreme meteorological conditions on influenza positivity risks

We further investigated the dynamic temporal variations in influenza positivity risks under extreme meteorological conditions (evaluating the 5^th^ and 95^th^ percentiles of each variable) to characterize type-specific vulnerability windows (Figures 5 A and B). Low temperature was associated with short-term increases in influenza A risk, with significant effects between lag 0.2 and 1 day and a peak at lag 0.2 days (RR = 1.55, 95% CI: 1.01–2.39). High temperature was not associated with influenza A. For influenza B, low temperature showed both immediate and delayed associations, with elevated risks at lag 0–5.6 days and 9.2–10.2 days. The strongest effect occurred at lag 0 days (RR = 1.56, 95% CI: 1.26–1.93). High temperature was also associated with influenza B risk at lag 4.2–7.2 days, peaking at lag 5.8 days (RR = 1.13, 95% CI: 1.04–1.23).

**Figure 5.**
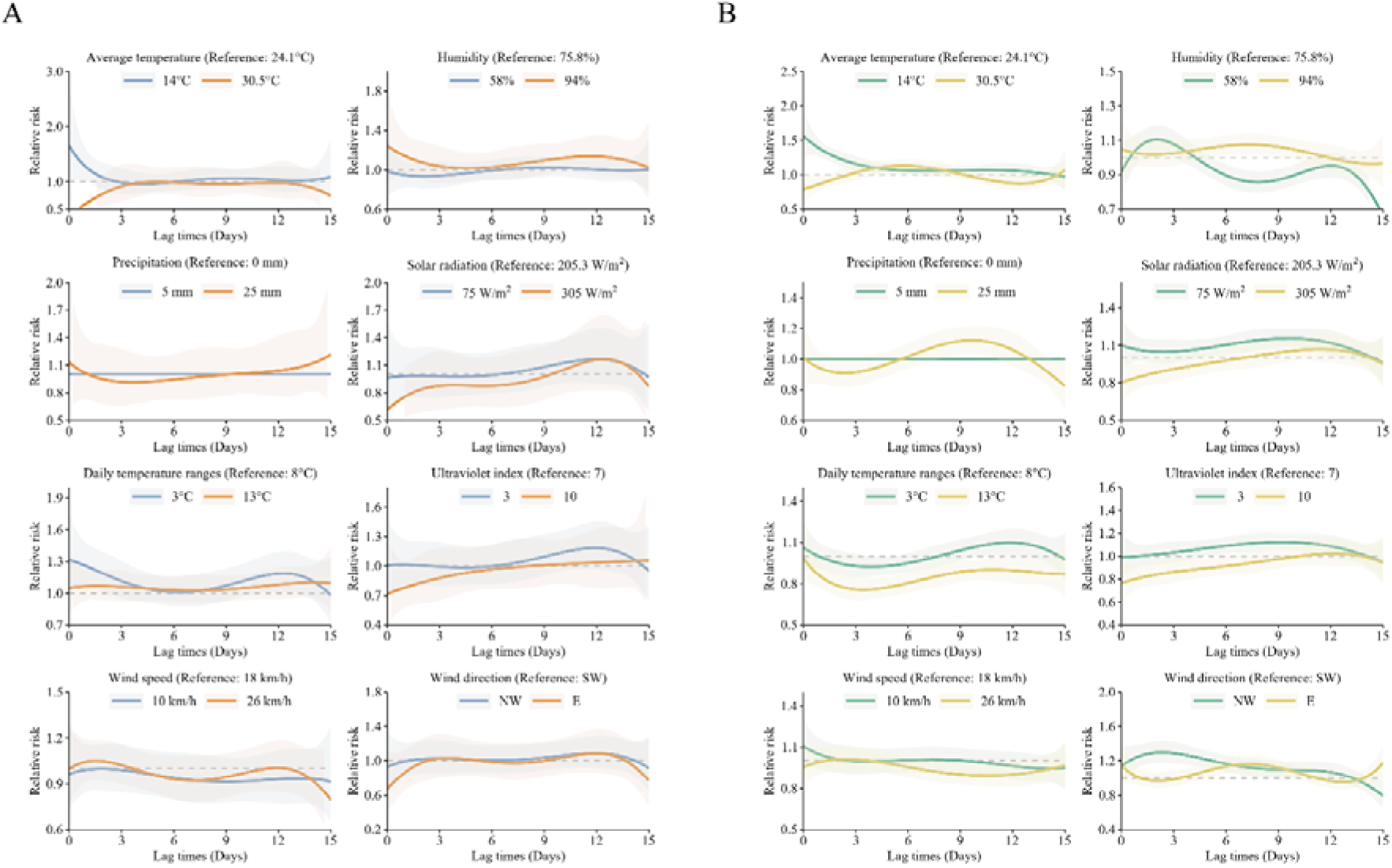
Lagged temporal effects of extreme meteorological conditions on Influenza A and B positivity rates. This figure visualizes the distribution of relative risks over a 15-day lag period following exposure to extreme meteorological conditions, derived from Distributed Lag Non-linear Model (DLNM). The analysis compares the temporal lag effects at the extreme high (95th percentile) and extreme low (5th percentile) values of selected weather variables for Influenza A (**A)** and Influenza B (**B)**. In each subplot, the x-axis represents the specific lag days (from day 0 to 15), and the y-axis shows the relative risk compared to the median reference condition. Solid orange lines (for Influenza A) and yellow lines (for Influenza B) represent extreme high-value conditions (e.g., extreme heat), whereas solid blue lines (for Influenza A) and green lines (for Influenza B) represent extreme low-value conditions (e.g., extreme cold). The shaded areas surrounding the curves represent the 95% confidence intervals. These lag-response curves elucidate the dynamic delay between specific extreme weather events and the subsequent peaks in influenza test positivity.

Under extreme humidity, neither very low (58%) nor very high (94%) humidity generated significant lagged risks for influenza A. However, for influenza B, low humidity posed a rapid risk (1.2–3 days, peak RR = 1.11 at day 2), while high humidity resulted in a delayed risk (5–9.2 days, peak RR = 1.08 at day 7.2). For precipitation, heavy rainfall (25 mm) uniquely impacted influenza B, inducing a lagged risk between 7.8 and 11 days (peak RR = 1.12 at day 9.6), with no significant effects observed for influenza A.

Low solar radiation and low UV index were consistently associated with delayed increases in influenza B risk. Low solar radiation increased influenza B risk from lag 4.6 to 12.6 days, peaking at lag 9.6 days (RR = 1.16, 95% CI: 1.08–1.23). Low UV index was associated with elevated influenza B risk from lag 5 to 11.8 days, peaking at lag 9.2 days (RR = 1.12, 95% CI: 1.05–1.20). No significant lagged associations were observed between these radiation-related variables and influenza A.

Extreme DTR showed limited but subtype-specific lagged effects. Low DTR was associated with influenza A risk at lag 0.2–2 days and 11–12.8 days, with peak estimates at lag 0.2 days (RR = 1.31, 95% CI: 1.02–1.66) and lag 12.2 days (RR = 1.19, 95% CI: 1.01–1.40). For influenza B, low DTR was associated with increased risk at lag 10.2–12.2 days, peaking at lag 11.8 days (RR = 1.10, 95% CI: 1.00–1.20). High DTR showed no significant association with either subtype. Wind-related effects were observed primarily for influenza B. Southwesterly winds were associated with increased influenza B risk from lag 0.4 to 10.4 days, peaking at lag 2.4 days (RR = 1.30, 95% CI: 1.17–1.44), while easterly winds were associated with risk from lag 5 to 9.8 days, peaking at lag 7.4 days (RR = 1.16, 95% CI: 1.07–1.26). Wind speed was not significantly associated with either influenza A or B.

Crucially, sensitivity analyses indicated that after removing the “weekly detection volumes” covariate, the reconstructed DLNM models remained structurally highly consistent with the original fully-adjusted models. This robust concordance confirms that the identified meteorological drivers represent intrinsic environmental triggers of viral transmission, independent of fluctuating surveillance intensity. Detailed methodological descriptions and results of the sensitivity analysis are presented in Supplementary Additional file 4.

### Influenza prediction using LSTM network

Building upon the previous analysis of the impact of meteorological factors on influenza dynamics, this study further aimed to construct a multifactorial influenza prediction network utilizing LSTM. To optimize the LSTM network, we employed Bayesian optimization through the Hyperopt framework, which allowed us to fine-tune key hyperparameters. This optimization process led to the identification of the optimal configurations for influenza A and B prediction algorithm (Table 1).

**Table 1:**
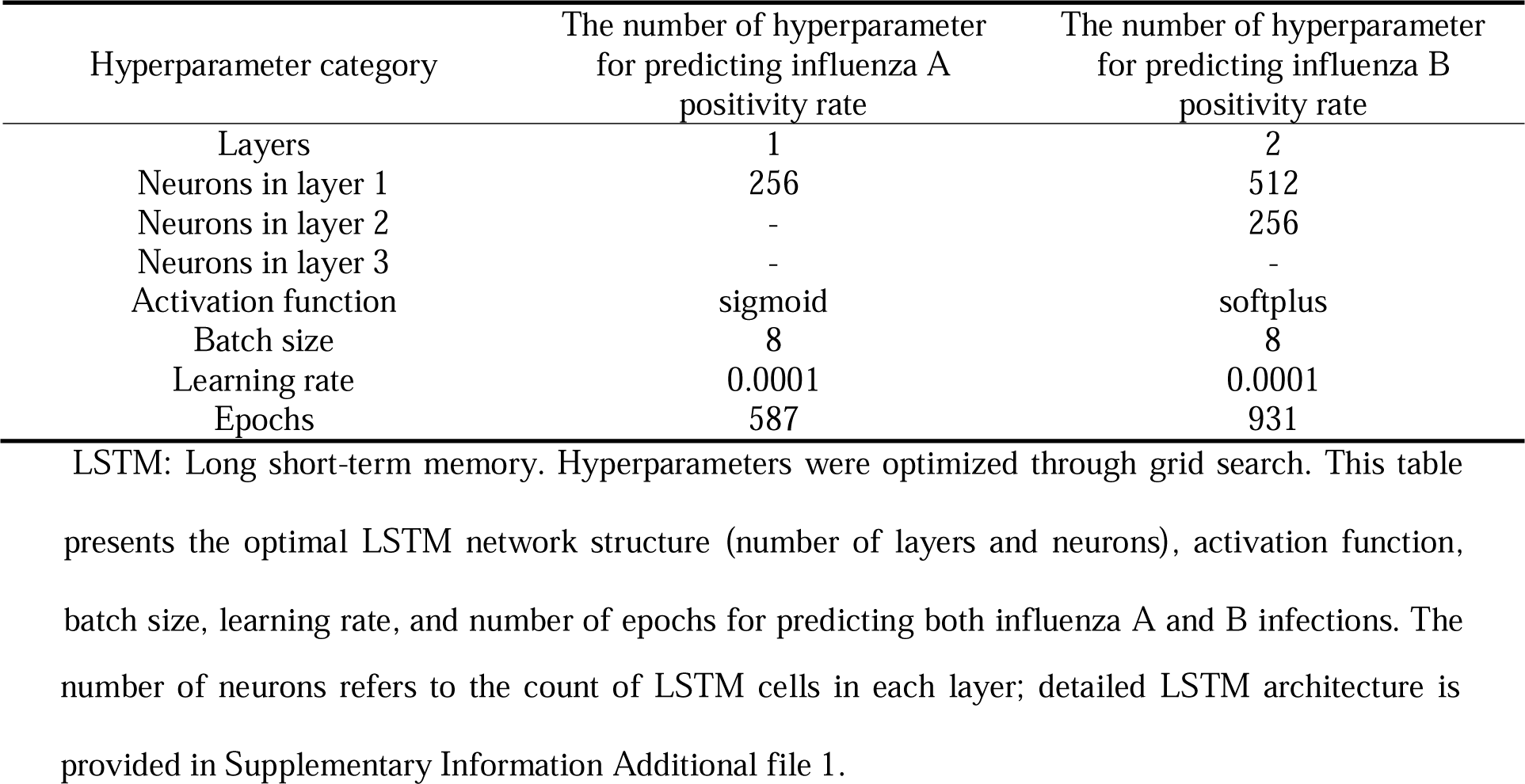
Optimal LSTM hyperparameters for predicting influenza A and B positivity rates.

For influenza A, the optimal LSTM network consisted of one hidden layer with 256 neurons, a learning rate of 0.0001, and a batch size of 8. In contrast, the influenza B LSTM network had two hidden layers with 512 and 256 neurons, a learning rate of 0.0001, and a batch size of 8. Leveraging these optimal hyperparameters, alongside a combination of meteorological data, weekly detection volumes, DOW, non-local population proportion, and mask-wearing stringency indices, we trained the LSTM network using historical data from 2018 to 2022. A 10% of the data were randomly selected as the validation set, while data from 2023 were subsequently used to test the network’s performance. The loss curves plotted during training revealed the convergence behavior of the LSTM. As shown in Figures 6 C and D, the training loss and validation loss for both the influenza A and B LSTM networks decreased progressively with increasing epochs, initially displaying a steep decline followed by stabilization. This pattern indicates that the algorithms effectively learned the characteristics of the data and converged to a stable solution. Furthermore, the close alignment between training and validation loss curves suggests that both LSTM networks for influenza A and B performed well and exhibited minimal overfitting, demonstrating strong learning capabilities and generalizability.

**Figure 6.**
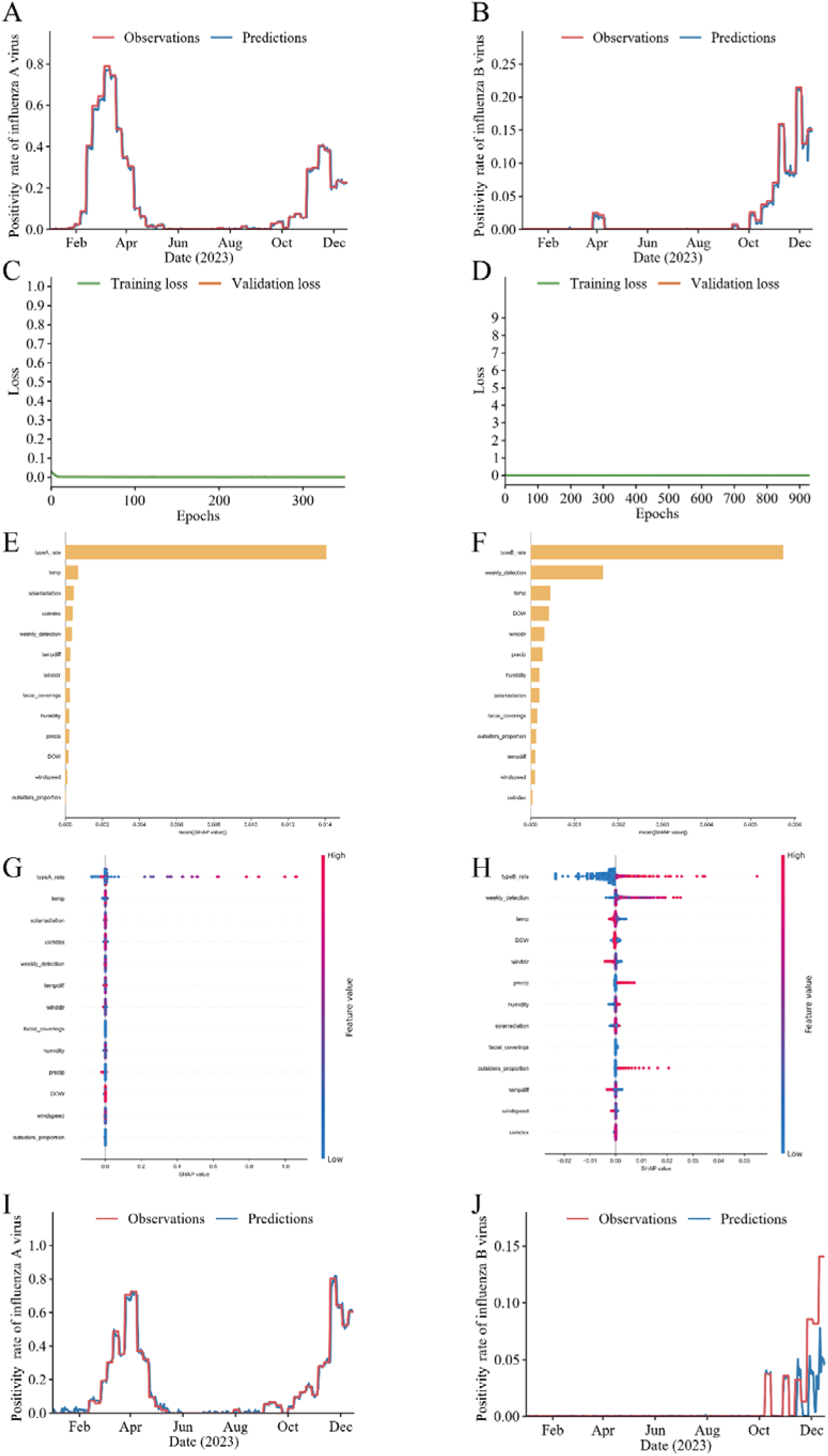
High-fidelity forecasting of the Influenza Positive Rate using the LSTM network and SHAP interpretability. This comprehensive panel visualizes the predictive performance and feature importance of the LSTM model configured to forecast the influenza positivity rate. **(C)** and **(D)** Training loss curves: These panels show the variation in loss values during the training of the LSTM network. The x-axis represents the number of training epochs, while the y-axis indicates the loss values. The smooth decline and stabilization of the loss curves indicate effective learning by the network, with no significant signs of overfitting. This is a crucial measure of the network’s stability and effectiveness throughout the training process. **(A)** and **(B)** Internal Out-of-Time validation (2023) comparing the LSTM-predicted positive rate (blue lines) against the observed positive rate (red lines) for Influenza A and B. The x-axis represents the date, and the y-axis reflects the positivity rates. This comparison visually demonstrates the network adeptly captures the non-linear viral rebound dynamics. **(E)** and **(F)** SHAP (SHapley Additive exPlanations) feature importance rankings. The bar charts quantify the mean absolute SHAP values, identifying historical subtype positivity rates (typeA_rate / typeB_rate), testing volume (weekly_detection), average temperature (temp), and mask-wearing stringency indices (facial_coverings) as paramount predictive drivers. The x-axis denotes the mean SHAP value, while the y-axis lists the variables. **(G)** and **(H)** SHAP summary plots illustrating the distribution of impacts for each feature on the model’s output, where color indicates feature value (red = high, blue = low). This graph provides a visual understanding of how and to what extent different features influence the influenza predictions, offering critical insights into the network’s decision-making process. (I) and (J) External spatial validation utilizing 2023 surveillance data from the neighboring city of Sanming, confirming the LSTM’s robust generalizability in forecasting independent epidemic curves. The date is plotted on the x-axis and the positivity rates are plotted on the y-axis.

Following adequate training, the LSTM networks were employed to predict influenza A and B positivity rates in Putian city for the year 2023. The results revealed a strong match between the predicted and observed values (Figures 6 A and B). The LSTM networks accurately captured both the timing and magnitude of these non-linear epidemic surges, including the two outbreak peaks of influenza A during February-March and November-December of 2023, as well as the peak of influenza B in November-December, demonstrating good predictive performance. The predictive performance of the LSTM networks was quantitatively assessed using metrics such as MAE, RMSE, MAPE, and SMAPE. For influenza A, the MAE was 0.009, RMSE was 0.035, MAPE was 0.158, and SMAPE was 0.521; for influenza B, the MAE was 0.002, RMSE was 0.011, MAPE was 0.17, and SMAPE was 0.484 (Table 2). These findings underscore the precision of the LSTM in characterizing and predicting influenza trends.

**Table 2:** Sensitivity analysis of the LSTM forecasting network evaluating the impact of epidemiological covariate ablation on predictive performance.

| Performance<br>evaluation<br>metric | Predicting Influenza A Positivity Rate |  |  | Predicting Influenza B Positivity Rate |  |  |
| --- | --- | --- | --- | --- | --- | --- |
|  | Full Model (All<br>variables<br>included) | Mask-wearing<br>index Removed | Weekly<br>detection<br>volumes<br>Removed | Full Model (All<br>variables<br>included) | Mask-wearing<br>index Removed | Weekly<br>detection<br>volumes<br>Removed |
| MAE | 0.009 | 0.012 | 0.011 | 0.002 | 0.003 | 0.004 |
| RMSE | 0.035 | 0.035 | 0.034 | 0.011 | 0.012 | 0.013 |
| MAPE | 0.158 | 0.247 | 0.203 | 0.170 | 0.135 | 0.120 |
| SMAPE | 0.521 | 0.846 | 0.791 | 0.484 | 0.857 | 1.450 |

Note: MAE (Mean Absolute Error) and RMSE (Root Mean Square Error) represent absolute predictive deviations. MAPE (Mean Absolute Percentage Error) and SMAPE (Symmetric Mean Absolute Percentage Error) assess relative accuracy. The full model incorporates meteorological variables alongside epidemiological covariates (mask-wearing stringency indices, weekly detection volumes, DOW, and migrant population proportion). Ablated models demonstrate the degradation in predictive accuracy upon the targeted removal of specific covariates, highlighting their necessity in adjusting for epidemiological non-stationarity.

To quantify the contributions of epidemiological and behavioral covariates to the network’s handling of pandemic non-stationarity, we conducted a covariate-ablation sensitivity analysis (Table 2). The LSTM networks were retrained sequentially by removing mask-wearing stringency indices and weekly detection volumes, while maintaining identical Bayesian-optimized hyperparameters. For influenza A, the exclusion of mask-wearing stringency indices resulted in a pronounced degradation of predictive performance compared to the full model: MAE, MAPE, and SMAPE increased by 33.3%, 56.3%, and 62.4%, respectively. The removal of weekly detection volumes similarly degraded performance, increasing MAE by 22.2% and SMAPE by 51.8%. This systemic shift in absolute errors (MAE) and the amplification of relative errors (SMAPE) clearly indicate that LSTM actively leverages mask mandates and testing intensities to capture the relative trends of influenza A. For influenza B, the ablation of covariates revealed an even higher dependency on the surveillance context. Removing weekly detection volumes proved highly detrimental, triggering a 100% surge in MAE, an 18.2% increase in RMSE, and a drastic 199.6% spike in SMAPE. This confirms that testing volumes act as a critical constraint variable for forecasting influenza B. Omitting mask-wearing indices similarly increased MAE by 50% and SMAPE by 77.1%.

Notably, a divergence was observed in the MAPE metric for influenza B, which paradoxically decreased (by 20.1% and 29.4%) upon the removal of these two covariates. This phenomenon is a well-documented mathematical artifact of the MAPE calculation, which utilizes the true observed value as the denominator. Given the extremely low baseline positivity rate of influenza B during certain periods, infinitesimal absolute deviations are asymmetrically magnified into massive percentage errors, causing MAPE to become highly unstable and misleading. In contrast, SMAPE, which utilizes the mean of the predicted and actual values in the denominator, effectively circumvents this asymmetric “low base-rate effect”. The profound deterioration of SMAPE across all ablated models corroborates the positive, stabilizing contribution of NPI and testing-volume covariates. These findings validate that our unified LSTM architecture does not merely rely on meteorological inputs, but effectively contextualizes epidemiological regime shifts to optimize forecasting precision.

To rigorously evaluate the spatial generalizability of the LSTM framework, we conducted an external validation utilizing concurrent 2023 surveillance and meteorological data from Sanming city. The network demonstrated robust transferability, achieving an MAE of 0.015, RMSE of 0.044, MAPE of 0.176, and SMAPE of 0.674 for influenza A; and an MAE of 0.005, RMSE of 0.018, MAPE of 0.487, and SMAPE of 1.792 for influenza B. As illustrated in Figures 6 I and J, the predicted trajectories successfully mirrored the actual observed incidence patterns in the new region, confirming the framework’s operational stability beyond its internal validation domain.

To benchmark the predictive value added by the LSTM architecture, we constructed two baseline models using identical training (2018–2022) and validation (2023) positivity-rate datasets, inclusive of all contextual covariates: the multivariate ARIMA model and the XGBoost gradient-boosting model. The LSTM model demonstrated decisive superiority over both baselines across all evaluated metrics (Supplementary Table S3). For influenza A, the LSTM achieved an MAE of 0.009 and SMAPE of 0.521, compared with the ARIMA’s MAE of 0.136 (SMAPE 1.212) and XGBoost’s MAE of 0.138 (SMAPE 1.081), representing approximately 15-fold error reductions relative to both benchmarks. For influenza B, the LSTM yielded an MAE of 0.002 (SMAPE 0.484), compared with 0.049 (SMAPE 0.810) for ARIMA and 0.070 (SMAPE 0.737) for XGBoost, approximately 25- to 35-fold reductions. Notably, ARIMA and XGBoost produced errors of comparable magnitude despite their disparate assumptions regarding linearity, suggesting that the LSTM’s advantage derives not from non-linear modeling capacity per se, but from architectural properties specific to recurrent sequential processing.

Beyond global error metrics, visual comparison of forecasting trajectories (Figure 6; Supplementary Figures S5 and S6) reveals critical behavioral disparities. For influenza A, both the linear ARIMA model and the non-linear XGBoost model generated conservatively smoothed forecasts that entirely failed to capture the sudden, explosive peaks of the 2023 post-restriction viral rebound. For influenza B, ARIMA produced continuous spurious fluctuations during non-epidemic periods (when true positivity was near zero), likely overreacting to covariate variations, while XGBoost generated largely flat trajectories that missed the mid-year outbreak peak. In stark contrast, the LSTM’s recurrent gating mechanisms successfully filtered out covariate noise during low-transmission periods while accurately tracking extreme epidemiological phase transitions, demonstrating a qualitative advantage in handling non-stationary regime shifts.

To open the computational “black box” and elucidate the predictive logic driving the LSTM networks, we conducted a SHAP analysis. The global feature importance rankings (Figures 6 E and F) revealed that the endogenous autoregressive component, historical influenza positivity rates, was the overwhelmingly dominant predictor for both subtypes. Beyond historical rates, the networks exhibited distinct, subtype-specific dependencies on meteorological and surveillance covariates. For influenza A (Figure 6 E), ambient temperature emerged as the second most impactful predictor, followed by solar radiation, the UV index, and weekly detection volumes. Conversely, for influenza B (Figure 6 F), weekly detection volumes emerged as the second most critical feature overall, followed by temperature and DOW. Crucially, these feature hierarchies confirm that the deep learning algorithm did not merely memorize spurious, weather-driven testing surges. Rather, it autonomously recognized the profound confounding impact of surveillance intensity bias. By assigning high predictive weight to weekly detection volumes and behavioral covariates, the LSTM network dynamically calibrated its epidemiological forecasts, effectively contextualizing intrinsic meteorological triggers within the rapidly shifting pandemic-era surveillance landscape.

## Discussion

This study characterized the population-level relationship between meteorological conditions and influenza activity in Putian, a representative non-megacity subtropical coastal city in southeastern China, using time-series (2018–2023) of integrated surveillance and meteorological data. As an ecological investigation, it was designed to describe environment–influenza associations and to develop an interpretable short-horizon forecasting framework, rather than to infer individual-level causality. By combining DLNM-based association analysis with Bayesian-optimized, subtype-specific LSTM forecasting, and by explicitly incorporating covariates related to testing intensity and NPI stringency, this work addresses three persistent gaps in subtropical influenza research: the isolated application of inferential and predictive tools, the limited use of principled hyperparameter optimization in LSTM-based influenza models, and the incomplete accommodation of pandemic-era non-stationarity in climate-sensitive forecasting. The findings deliver subtype-specific insights into meteorology–influenza coupling and establish a methodologically rigorous foundation for future operational surveillance systems in subtropical settings.

Unlike temperate regions, subtropical areas exhibit year-round influenza circulation characterized by considerable epidemiological heterogeneity. A Hong Kong-based study reported that local influenza A typically peaks between January and April, whereas influenza B peaks from May to July (Kim, Park, and Lee 2020). A study from Okinawa, Japan, identified analogous trends, with influenza A peaking from December to March and influenza B from April to June (Iha et al. 2016). Our six-year dataset revealed that both viruses circulated year-round in Putian, with peaks concentrated in winter-spring and additional summer activity. Influenza A predominantly peaked between December and April, whereas influenza B outbreaks frequently occurred outside this conventional window, suggesting that extreme weather fluctuations, rather than mere seasonality, act as primary outbreak triggers (Dave and Lee 2019). The infection burden was concentrated among individuals under 18 years (73.12%), with male predominance in childhood and female predominance in adulthood, consistent with heightened exposure risk in school-aged children through close-contact group settings (Jackson et al. 2013). Notably, the significant suppression of influenza positivity observed in 2020 likely reflects the impact of strict NPIs and altered population contact networks implemented during the early COVID-19 period. Collectively, these patterns motivated the subsequent quantitative characterization of subtype-specific meteorological drivers.

The meteorology-influenza relationship remains a critical question in environmental epidemiology, with ambient temperature, relative humidity, precipitation, DTR, and wind parameters known to modulate virion survival, host respiratory susceptibility, and population contact behaviors (Polozov et al. 2008; Sooryanarain and Elankumaran 2015). Consistent with the intrinsic non-linearity of these relationships, Pearson correlation coefficients Figure S2) were uniformly weak (all *r* < 0.3), reinforcing the inadequacy of linear between influenza positivity rates and meteorological variables in our cohort (Supplementary approaches. DLNM cross-basis analysis, in contrast, revealed clear non-linear exposure–response and lag–response structures that differed markedly between subtypes.

Cumulative exposure–response patterns diverged notably between subtypes. Influenza A positivity was associated with warmer conditions between 15°C and 24°C, with the cumulative relative risk peaking at 15°C (RR = 3.01, 95% CI: 1.03–8.78). Conversely, the cumulative risk for influenza B progressively declined from 8°C to 24°C, peaking sharply at the colder threshold of 8°C (RR = 26.12, 95% CI: 7.79–87.61). This distinct thermal preference contrasts with findings from Shanghai, where both subtypes showed maximal cumulative risk near 1.4°C (Zhang et al. 2020). Enhanced transmission at lower temperatures is broadly consistent with greater envelope stability in cold air (Lowen and Steel 2014), impaired mucociliary clearance (Eccles 2002), and increased indoor confinement facilitating aerosol exchange (Liao, Chang, and Liang 2005). Conversely, elevated temperatures can also inadvertently increase transmission through reduced outdoor activity and greater reliance on closed-cycle air conditioning (Lofgren et al. 2007). The subtype-specific divergence may further reflect fundamental virological and immunological differences, including the relative stability of influenza A glycoproteins under warmer, humid conditions and a structural preference of influenza B for cooler, wetter environments (Marr et al. 2019). Immunologically, high ambient temperatures can impair adaptive immune responses, potentially facilitating influenza A infection (Moriyama and Ichinohe 2019), whereas cooler settings preserve innate responses, aiding the control of influenza B (Lofgren et al. 2007). Complex host-environment interactions may also suppress specific immune components, such as immunoglobulin M, inadvertently creating a permissive environment for influenza B transmission (Xu, Hu, and Tian 2017).

High relative humidity has been widely documented to enhance influenza risk by promoting viral transmission, as corroborated by multi-city studies in the Sichuan Basin (Zhou et al. 2022). Cellular models indicate that humidity influences respiratory droplets evaporation, altering solute concentration and influenza A virus viability (Yang, Elankumaran, and Marr 2012), while findings from Arizona indicate increased influenza risk during the rainy season (Soebiyanto, Adimi, and Kiang 2010). Consistent with these observations, our results showed that the cumulative risk for influenza B gradually escalated within a high-humidity range of 76% to 91%, peaking at 91% (RR = 1.52, 95% CI: 1.00–2.29). The cumulative risk for influenza B also progressively increased with decreasing solar radiation, peaking 10 W/m² (RR = 8.66, 95% CI: 2.14–35.00), suggesting that insufficient sunlight exposure may specifically promote influenza B transmission, a pattern absent for influenza A. Extensive literature supports that adequate solar radiation and subsequent UV exposure reduce airborne influenza viability and support host vitamin D synthesis and immune modulation (Cannell et al. 2006; Sagripanti and Lytle 2007). However, contradictory effects of UV and solar radiation on subtypes A and B reported in pediatric Vitamin D supplementation trials (Urashima et al. 2010) necessitate further mechanistic investigation into the interplay between radiant energy and specific viral lineages. Physical mechanical factors such as DTR and wind metrics also demonstrated nuanced impacts. Prior research indicates that DTR serves as a critical risk factor, with severe fluctuations (11–14°C) increasing infection risk by compromising respiratory epithelial barriers and modulating inflammatory mucus production (Park et al. 2020). Our analysis partially corroborated this, showing that narrower DTRs (1–8°C) heightened cumulative risks for both subtypes, potentially reflecting sustained susceptibility under chronically stable yet suboptimal thermal conditions (Cheng et al. 2014). Wind direction influenced only influenza B, while wind speed exerted no significant cumulative effect on either subtype, consistent with literature deeming wind speed a non-critical epidemiological variable (Ianevski et al. 2019).

Lag structures further distinguished the subtypes within the 15-day post-exposure window. Low average temperatures of 14°C were associated with a rapid 1-day lagged effect for influenza A, but induced a prolonged, bimodal lagged risk for influenza B (0–5 days and 9–10 days). Conversely, high averages of 30.5°C conferred a delayed risk for influenza B only at 4.2–7.2 days, a timeline closely mirroring Wang et al.’s reported 7.3-day lag effect for influenza B at 28°C (Wang et al. 2023). Extreme high precipitation (25 mm) induced a delayed risk for influenza B at 7.8–11 days, consistent with meta-analyses confirming that extreme rainfall events in the subtropics trigger lagged infectious disease surges (Aune, Davis, and Smith 2021). Notably, our data revealed a profound paradox regarding diurnal temperature variation. While a high DTR (13°C) exhibited significant cumulative relative risk, it produced no distinct lagged risk peaks. In stark contrast, a low DTR (3°C) triggered pronounced lagged risks for both influenza A (0–2 and 11–12.8 days) and influenza B (10.2–12.2 days). The intrinsic physiological mechanisms underlying this divergence, where broad temperature swings drive overall cumulative burden, but narrow temperature bands dictate specific temporal outbreak windows, warrant deeper investigation. Overall, these results identify the 1-to-2-week window following specific meteorological anomalies as an operationally meaningful period for anticipatory action, while emphasizing that thresholds and lag structures require local calibration. The fixed 15-day maximum lag adopted here, though consistent with prior literature, may not capture all longer-range environmental effects; systematic sensitivity analyses varying the lag window would be a valuable extension.

A central methodological feature of our framework is the deliberate division of labor between the DLNM and LSTM components. Multivariate DLNMs efficiently expose transparent, lag-resolved main-effect surfaces for each meteorological variable, but cannot accommodate high-dimensional interactions among meteorological, autoregressive, and socio-behavioral factors without parameter inflation and severe multicollinearity. The LSTM stage was therefore not intended to replace DLNM inference, but to complement it by learning joint non-linear structure across concurrent covariates. Furthermore, extensive environmental inputs inevitably introduce severe collinearity, such as the strongly correlated solar radiation and UV index. While traditional multivariate models are highly vulnerable to such overlapping variances, the recurrent, weighted representation learned by the LSTM is comparatively tolerant of such redundancy, allowing broader covariate integration than in previous efforts (Zhu et al. 2022). Crucially, this LSTM stage explicitly incorporates weekly detection volumes, mask-wearing stringency indices, non-local population proportion, and DOW effects alongside meteorological inputs, and uses influenza positivity rates rather than absolute case counts as the modeling endpoint. Together, these design choices are intended to mitigate, rather than fully eliminate, the surveillance-related biases that can distort count-based forecasting during periods of fluctuating testing intensity.

To maximize this architectural advantage and ensure mathematical robustness, we conducted refined hyperparameter optimizations using the Hyperopt framework (Li, Zhang, and Ma 2023). By employing a Bayesian strategy based on Tree-based Parzen Estimators (TPE) (Hanifi et al. 2022), our approach efficiently navigated the complex, high-dimensional hyperparameter space. Unlike traditional heuristic or manual grid search methods, this Bayesian approach systematically optimized the learning rate, network depth, and activation functions to strike an optimal balance between model fitting capacity and the prevention of overfitting (Hanifi, Cammarono, and Zare-Behtash 2024). To our knowledge, this represents a novel application of Hyperopt-based tuning for an epidemiological LSTM forecasting network, ensuring that our algorithm’s superior performance is grounded in mathematically rigorous architectural configuration rather than arbitrary parameter selection.

The optimized LSTM algorithm demonstrated strong predictive performance, with loss curves for both subtype-specific networks exhibiting favourable convergence, consistent with prior work (Du et al. 2023). To rigorously isolate the predictive contribution attributable to the LSTM’s recurrent architecture, we benchmarked it against two covariate-matched baselines representing distinct methodological paradigms: the multivariate ARIMA model and the XGBoost gradient-boosting ensemble. This design controls simultaneously for linearity (ARIMA→LSTM contrast) and for non-linearity without recurrent memory (XGBoost→LSTM contrast), allowing us to attribute observed performance gains to specific architectural inductive biases rather than to informational asymmetry or model non-linearity in general. The LSTM substantially outperformed both baselines on the 2023 validation window. For influenza A, it achieved an MAE of 0.009, compared with 0.136 for ARIMA and 0.138 for XGBoost, approximately 15-fold reductions. For influenza B, the LSTM yielded an MAE of 0.002, versus 0.049 for ARIMA and 0.070 for XGBoost, 25- to 35-fold reductions. Critically, ARIMA and XGBoost produced errors of comparable magnitude despite their disparate assumptions regarding linearity, and both systematically under-predicted the explosive 2023 post-NPI rebound for influenza A while generating flat or spurious trajectories for influenza B (Supplementary Figures S5–S6). That parallel failure indicates the LSTM’s advantage under pandemic-era non-stationarity derives not from non-linearity per se, but from four architectural properties intrinsic to recurrent gated networks yet absent in tree ensembles: (i) threshold-based, sequence-insensitive splits fail to encode present–past dynamics; (ii) XGBoost lacks forget–input–output gates to propagate and re-weight historical states across time lags; (iii) gradient-boosted trees assume near-independence, contradicting the pronounced temporal autocorrelation in epidemiological time series; (iv) ARIMA and LSTM are sequential models with different functional forms, whereas XGBoost is non-sequential. The joint failure of ARIMA and XGBoost, despite differing non-linear treatment, isolates recurrent sequential memory as the critical architectural feature for forecasting under non-stationarity. We emphasize that this interpretation applies specifically to the present non-stationary influenza forecasting task and does not constitute a general dismissal of gradient-boosted ensembles, which retain state-of-the-art performance across many structured prediction domains. Rather, it highlights that for surveillance time series exhibiting pronounced temporal dependencies and abrupt regime shifts, such as the 2023 post-NPI rebound, explicit sequential memory becomes functionally essential. This mechanistic reading, together with the LSTM’s comparative edge over previously reported ARIMA-based (Li et al. 2024) and LSTM-based influenza prediction models (Zhu et al. 2022), positions our framework as a substantive methodological advance in predictive modeling for climate-sensitive diseases.

External evaluation using independent Sanming CDC data yielded MAE values of 0.0152 for influenza A and 0.0054 for influenza B, providing preliminary evidence of model portability within the southeastern Chinese subtropical context, though this rests on a single external city and should not be interpreted as broad spatial generalizability. SHAP analysis (Figure 6 E–F) further enhanced LSTM interpretability, identifying historical influenza positivity rates, weekly detection volumes, temperature, and solar radiation as the most highly weighted predictive features, a pattern consistent with temperature- and radiation-mediated influenza virus inactivation mechanisms (Sagripanti and Lytle 2007), and corroborating the network’s autonomous capacity to calibrate predictions against fluctuating surveillance intensities. While SHAP’s underlying assumption of feature independence may not fully capture temporal interactions within the LSTM network, treating each meteorological variable’s lagged sequence as a unified feature partially preserves temporal dependencies. Future work could explore temporal SHAP extensions or alternative interpretability methods designed for sequential data.

Beyond its forecasting accuracy, the DLNM-LSTM framework offers several potential contributions for subtropical public health practice. By delineating subtype-specific meteorological thresholds, characterizing their lagged temporal effects, and dynamically integrating behavioral and surveillance covariates, it enables the anticipation of high-risk transmission windows 1 to 2 weeks in advance of clinical signal emergence. Such foresight can inform pre-emptive vaccination campaigns for vulnerable populations, optimize antiviral stockpiling and healthcare resource allocation, and intensify surveillance during predicted risk windows to accelerate case detection and containment. The underlying dual-stage architecture is also methodologically extensible to other climate-sensitive infectious diseases, including dengue and hand-foot-and-mouth disease, in which non-linear environmental coupling and lagged effects similarly dominate transmission dynamics. In a warming climate, where influenza seasonality may shift in response to evolving weather regimes, the framework’s modular design permits periodic recalibration to maintain predictive validity. Importantly, however, this framework should be regarded as a methodological foundation for future operational developments rather than as a deployable early-warning system: routine use in public health practice would require prospective recalibration, integration with operational surveillance infrastructure, and additional validation beyond the scope of the present study.

This study has several limitations that should be considered when interpreting the findings. Firstly, our primary analysis relies on influenza surveillance data collected from seven sentinel hospitals in Putian, which inherently captures only a fraction of all influenza cases occurring in the broader community. Although employing positivity rates as the primary outcome substantially mitigates the surveillance bias inherent in count-based analyses, residual selection effects may persist if testing patterns shift differentially across demographic subgroups with systematically divergent positivity profiles. Our inclusion of weekly detection volumes as an explicit LSTM covariate, complemented by parallel DLNM sensitivity analyses validating the independence of meteorological effects from testing volumes, were designed to characterize and partially account for this residual bias, but cannot fully eliminate it. Consequently, positivity rates likely underestimate the true burden of community influenza infection. Although the surveillance infrastructure in Putian remained uniform and uninterrupted throughout the 2018–2023 timeline, mitigating measurement bias and temporal confounding risks, the transferability of our findings to other global subtropical regions with different socioeconomic structures, healthcare systems, or population behaviors requires cautious, region-specific calibration. Accordingly, our framework should be strictly interpreted as a high-fidelity tool designed to forecast the observable public health surveillance signal, which is the most operationally relevant target for public health agencies, rather than for estimating unobserved, absolute community disease burden. Future research should aim to integrate broader data sources, such as community-level surveillance or syndromic data streams where available, and pursue multi-centre studies across diverse global subtropical regions to disentangle core meteorological determinants that transcend geographical boundaries from those requiring local adaptation.

Secondly, the interpretation of our framework’s predictive performance during the 2023 validation period requires careful epidemiological and methodological contextualization. The year of 2023 represented an anomalous, post-restriction “rebound” period characterized by rapid NPI relaxation and the release of accumulated population-level immunity debt, resulting in an atypical influenza surge that exceeded pre-pandemic peaks. The framework’s high accuracy across this period should therefore be interpreted as evidence of algorithmic agility and adaptive capacity during a highly volatile transitional phase, rather than as definitive proof of long-term predictive validity under a stabilized post-2024 epidemiological regime. Methodologically, while our strict chronological OOT data partitioning prevented temporal information leakage, a critical requirement for LSTM integrity, the reliance on a single, fixed chronological split point (December 31, 2022) intrinsically limits our evaluation to one specific structural break. This fixed-split approach may not exhaustively probe the DLNM-LSTM framework’s resilience against all forms of future epidemiological non-stationarity. Consequently, naive extrapolation of the reported 2023 performance metrics to future surveillance years should be avoided absent prospective recalibration. Future studies should consider employing expanding-window or rolling-origin cross-validation frameworks to provide a more continuous characterization of algorithmic robustness. Continuous integration of accumulating 2024 and 2025 data, combined with adaptive learning architectures capable of detecting regime shifts in real time, will be essential before any operational deployment of this, or similar forecasting frameworks, for routine public health surveillance.

Thirdly, beyond the surveillance coverage limitation noted above, the aggregate-level nature of the available data further constrained our ability to adjust for individual-level confounders, including personal vaccination status, detailed comorbidities, healthcare-seeking behavior, and socioeconomic status. This limitation was particularly exacerbated by the profound epidemiological disruptions during the COVID-19 pandemic, where raw numbers of confirmed cases became heavily confounded by surveillance intensity (i.e., fluctuating testing volumes) rather than solely reflecting underlying viral transmission. Our adoption of influenza positivity rates as the primary modeling endpoint and incorporation of weekly detection volumes, face-covering stringency, and non-local population proportion as dynamic covariates, rigorously mitigated these aggregate-level biases and linked our methodological design directly to the forecasting outcomes. As unequivocally demonstrated by our covariate-ablation sensitivity analyses (Table 2), failing to account for mask mandates and testing volumes leads to severe, mathematically predictable deviations in absolute forecasting accuracy. Furthermore, while daily case counts in a single city can occasionally be small, sporadic, and driven by external importations, our incorporation of non-local population proportion effectively adjusted for these localized importation risks. Consequently, although our findings characterize population-level associations between meteorological factors and influenza activity, they should not be interpreted as evidence of micro-level causal mechanisms at the individual patient level. Ultimately, the DLNM-LSTM framework’s robust performance across the non-stationary transition out of NPI policies highlights the absolute necessity of integrating behavioral and virological baseline metrics into future climate-driven predictive surveillance systems.

Fourthly, our benchmarking strategy was deliberately structured as a paradigmatically layered three-way comparison (linear-sequential ARIMA, non-linear non-sequential XGBoost, non-linear sequential LSTM) rather than an exhaustive algorithmic survey. This design prioritized methodological clarity, isolating the contribution of recurrent sequential inductive bias, over horizontal coverage. Nonetheless, our evaluation does not extend to Transformer-based attention architectures, nor to hybrid ensemble strategies (e.g., LSTM–XGBoost stacking or multi-model Bayesian model averaging) that may offer complementary strengths. Systematic benchmarking against these emerging architectures, alongside prospective recalibration on additional subtropical surveillance streams and operational stress-testing under real-time data latency, will be essential next steps as the framework evolves toward operational deployment.

Finally, it is imperative to explicitly delineate the generalizability of our forecasting framework within geographic and epidemiological contexts. The successful external validation in Sanming provides direct empirical evidence that our LSTM architecture and the identified meteorological thresholds are robustly transferable across the subtropical southeastern Chinese context, which shares comparable monsoon climates, year-round influenza circulation, and standardized public health intervention frameworks. However, we explicitly define the boundaries of this transferability. The specific meteorological coefficients, lag structure, and predictive parameters derived in this study should not be indiscriminately extrapolated to other regions, even within subtropical latitudes, where climatic regimes (e.g., non-monsoon subtropics) or public health contexts (e.g., higher baseline influenza vaccination coverage or distinct NPI strategies) differ substantially. In such disparate settings, directly applying our pretrained model weights may yield substantial systematic biases. While the underlying dual-modeling architecture remains methodologically transferable, its parameterization requires rigorous local recalibration using region-specific surveillance data. Acknowledging these boundaries ensures that the framework can be deployed safely and appropriately to realize targeted, climate-sensitive infectious disease forecasts.

In conclusion, this study elucidates the distinct, non-linear meteorological drivers of influenza A and B transmission in a subtropical Chinese urban setting through an integrated dual-stage DLNM-LSTM framework. By adopting influenza positivity rates as the primary outcome and integrating socio-behavioral covariates, including mask-wearing stringency indices, weekly detection volumes, and non-local population proportion indicators, our approach mitigates surveillance-related biases and accommodates pandemic-era non-stationarity. It shows lower forecast errors than covariate-matched baselines, both linear autoregressive (ARIMA) and non-linear gradient-boosting (XGBoost), and provides preliminary evidence of portability within southeastern subtropical China, combining the interpretability of distributed lag modeling with the flexibility and sequential inductive bias of deep learning. The framework offers an interpretable, climate-informed methodological foundation for future operational surveillance developments in subtropical settings.

## Data Availability

The influenza surveillance data and meteorological records utilized in this study, as well as the code written for constructing the DLNM models and LSTM algorithms, are available upon reasonable request to the corresponding authors X-W. J. and M-J. Z. All data are managed in accordance with the ethical standards and data management policies of the affiliated institutions. Requests for data access will be reviewed and fulfilled in compliance with these policies and regulations.
The computational codes used in this study, including those for the distributed lag non-linear models (DLNM) and the Bayesian-optimized Long Short-Term Memory (LSTM) neural network implementations, were written in R and Python. All custom scripts used for data analysis, model development, and visualization are available upon reasonable request from the corresponding authors X-W. J. and M-J. Z. The codes include data preprocessing, model training, validation procedures, and the generation of prediction results. Researchers interested in reproducing or building upon our methodology may contact the corresponding authors with specific requirements regarding the code components needed for their research purposes.

## Acknowledgements

We acknowledge the invaluable contribution of the Putian CDC and its affiliated sentinel hospitals to this study. We express our deep respect and gratitude to the dedication of all participating healthcare workers, disease prevention personnel, clinical medical staff, and laboratory technicians involved in case screening, data recording, sample collection, and laboratory testing for their significant efforts and contributions to this study.

## Author Contributions

Conceptualisation: X-W. J., L-M. L., X. H., S. X., and L. X; Data curation: M-J. Z., J-L. T., Z-S. W., Y-X. L., J-L. C., J-J. H., J. Q., and H-Y. P; Formal analysis: L. X., M-J. Z., J-L. T., Z-S. W., Y-X. L., Z-Q. X., S. X., and X. H; Funding acquisition: L-M. L. and X-W. J; Investigation: M-J. Z., Y-X. L., J. Q., J-J. H., J-L. C., H-Y. P., and Z-F. R; Methodology: J-L. T., Z-S. W., L. X., X-W. J., X. H., S. X., and M-J. Z; Software: J. Q., H-Y. P., J-L. T., Z-S. W., and Z-F. R; Project administration: X-W. J., L-M. L., and X. H; Resource: X-W. J, L-M. L., S. X., and X. H; Supervision: X-W. J., L-M. L., M-J. Z., and Y-X. L.; Validation: L. X., Y-X. L., J-L. T., Z-S. W., Z-Q. X., X. H., and M-J. Z; Visualisation: L. X., J-L. T., Z-S. W., J-L. C., J. Q., J-J. H., and Z-F. R; Writing-original draft: L. X., and J-L. T; Writing-review & editing: L. X., X-W. J., L-M. L., S. X., and X. H. All authors have reviewed and approved the final version of the manuscript.

## Availability of Data Statement

The influenza surveillance data and meteorological records utilized in this study, as well as the code written for constructing the DLNM models and LSTM algorithms, are available upon reasonable request to the corresponding authors X-W. J. and M-J. Z. All data are managed in accordance with the ethical standards and data management policies of the affiliated institutions. Requests for data access will be reviewed and fulfilled in compliance with these policies and regulations.

## Code Availability Statement

The computational codes used in this study, including those for the distributed lag non-linear models (DLNM) and the Bayesian-optimized Long Short-Term Memory (LSTM) neural network implementations, were written in R and Python. All custom scripts used for data analysis, model development, and visualization are available upon reasonable request from the corresponding authors X-W. J. and M-J. Z. The codes include data preprocessing, model training, validation procedures, and the generation of prediction results. Researchers interested in reproducing or building upon our methodology may contact the corresponding authors with specific requirements regarding the code components needed for their research purposes.

## Supplementary information

**Table S1: Descriptive Statistics of Meteorological Factors in Putian City (2018-2023).**

**Table S2: QAIC Values for DLNM Models Across Degrees of Freedom Combinations.**

**Table S3. Comparative predictive performance of LSTM, multivariate ARIMA, and XGBoost on the 2023 validation set.**

**Figure S1: Detailed Structure of the Long Short-Term Memory (LSTM) Cell.**

**Figure S2: Correlation Heatmap and Scatterplot Matrix Illustrating Pairwise Associations and Potential Collinearity among Influenza Indicators and Meteorological Variables.**

**Figure S3. Sensitivity Analysis of the Cumulative Risk of Meteorological Factors on Influenza A and B Positivity Rates.**

**Figure S4. Sensitivity Analysis of the Lagged Temporal Effects of Extreme Meteorological Conditions on Influenza A and B Positivity Rates.**

**Figure S5. The Actual Values of Influenza A and B Compared to the Predicted Values from the Covariate-Adjusted ARIMA Model.**

**Figure S6. Observed versus XGBoost-predicted weekly positivity rates for (A) Influenza A and (B) Influenza B in Putian City, 2023.**

## REFERENCES

Absar, N., N. Uddin, M. U. Khandaker, and H. Ullah. 2022. ’The efficacy of deep learning based LSTM model in forecasting the outbreak of contagious diseases’, Infect Dis Model, 7: 170–83.

Aune, K. T., M. F. Davis, and G. S. Smith. 2021. ’Extreme Precipitation Events and Infectious Disease Risk: A Scoping Review and Framework for Infectious Respiratory Viruses’, Int J Environ Res Public Health, 19.

Bloom-Feshbach, K., W. J. Alonso, V. Charu, J. Tamerius, L. Simonsen, M. A. Miller, and C. Viboud. 2013. ’Latitudinal variations in seasonal activity of influenza and respiratory syncytial virus (RSV): a global comparative review’, PLoS One, 8: e54445.

Cannell, J. J., R. Vieth, J. C. Umhau, M. F. Holick, W. B. Grant, S. Madronich, C. F. Garland, and E. Giovannucci. 2006. ’Epidemic influenza and vitamin D’, Epidemiol Infect, 134: 1129–40.

Cheng, J., Z. Xu, R. Zhu, X. Wang, L. Jin, J. Song, and H. Su. 2014. ’Impact of diurnal temperature range on human health: a systematic review’, Int J Biometeorol, 58: 2011–24.

Chicco, D., M. J. Warrens, and G. Jurman. 2021. ’The coefficient of determination R-squared is more informative than SMAPE, MAE, MAPE, MSE and RMSE in regression analysis evaluation’, PeerJ Comput Sci, 7: e623.

Dave, K., and P. C. Lee. 2019. ’Global Geographical and Temporal Patterns of Seasonal Influenza and Associated Climatic Factors’, Epidemiol Rev, 41: 51–68.

Deyle, E. R., M. C. Maher, R. D. Hernandez, S. Basu, and G. Sugihara. 2016. ’Global environmental drivers of influenza’, Proc Natl Acad Sci U S A, 113: 13081–86.

Du, X., L. Si, P. Li, and Z. Yun. 2023. ’A method for detecting the quality of cotton seeds based on an improved ResNet50 model’, PLoS One, 18: e0273057.

Eccles, R. 2002. ’An explanation for the seasonality of acute upper respiratory tract viral infections’, Acta Otolaryngol, 122: 183–91.

Gasparrini, A. 2011. ’Distributed Lag Linear and Non-Linear Models in R: The Package dlnm’, J Stat Softw, 43: 1–20.

Gasparrini, A., B. Armstrong, and M. G. Kenward. 2010. ’Distributed lag non-linear models’, Stat Med, 29: 2224–34.

Groves, H. E., P. P. Piché-Renaud, A. Peci, D. S. Farrar, S. Buckrell, C. Bancej, C. Sevenhuysen, A. Campigotto, J. B. Gubbay, and S. K. Morris. 2021. ’The impact of the COVID-19 pandemic on influenza, respiratory syncytial virus, and other seasonal respiratory virus circulation in Canada: A population-based study’, Lancet Reg Health Am, 1: 100015.

Guo, Q., Z. Dong, W. Zeng, W. Ma, D. Zhao, X. Sun, S. Gong, J. Xiao, T. Li, and W. Hu. 2019. ’The effects of meteorological factors on influenza among children in Guangzhou, China’, Influenza Other Respir Viruses, 13: 166–75.

Hanifi, Shahram, Andrea Cammarono, and Hossein Zare-Behtash. 2024. ’Advanced hyperparameter optimization of deep learning models for wind power prediction’, Renewable Energy, 221: 119700.

Hanifi, Shahram, Saeid Lotfian, Hossein Zare-Behtash, and Andrea Cammarano. 2022. ’Offshore Wind Power Forecasting—A New Hyperparameter Optimisation Algorithm for Deep Learning Models’, Energies, 15: 6919.

Ianevski, A., E. Zusinaite, N. Shtaida, H. Kallio-Kokko, M. Valkonen, A. Kantele, K. Telling, I. Lutsar, P. Letjuka, N. Metelitsa, V. Oksenych, U. Dumpis, A. Vitkauskiene, K. Stašaitis, C. Öhrmalm, K. Bondeson, A. Bergqvist, R. J. Cox, T. Tenson, A. Merits, and D. E. Kainov. 2019. ’Low Temperature and Low UV Indexes Correlated with Peaks of Influenza Virus Activity in Northern Europe during 2010ß2018’, Viruses, 11.

Iha, Y., T. Kinjo, G. Parrott, F. Higa, H. Mori, and J. Fujita. 2016. ’Comparative epidemiology of influenza A and B viral infection in a subtropical region: a 7-year surveillance in Okinawa, Japan’, BMC Infect Dis, 16: 650.

Iuliano, A. D., K. M. Roguski, H. H. Chang, D. J. Muscatello, R. Palekar, S. Tempia, C. Cohen, J. M. Gran, D. Schanzer, B. J. Cowling, P. Wu, J. Kyncl, L. W. Ang, M. Park, M. Redlberger-Fritz, H. Yu, L. Espenhain, A. Krishnan, G. Emukule, L. van Asten, S. Pereira da Silva, S. Aungkulanon, U. Buchholz, M. A. Widdowson, and J. S. Bresee. 2018. ’Estimates of global seasonal influenza-associated respiratory mortality: a modelling study’, Lancet, 391: 1285–300.

Jackson, C., E. Vynnycky, J. Hawker, B. Olowokure, and P. Mangtani. 2013. ’School closures and influenza: systematic review of epidemiological studies’, BMJ Open, 3.

Javanian, M., M. Barary, S. Ghebrehewet, V. Koppolu, V. Vasigala, and S. Ebrahimpour. 2021. ’A brief review of influenza virus infection’, J Med Virol, 93: 4638–46.

Kim, B. I., O. Park, and S. Lee. 2020. ’Comparison of influenza surveillance data from the Republic of Korea, selected northern hemisphere countries and ß Hong Kong Special Administrative Region SAR (China) from 2012 to 2017’, Western Pac Surveill Response J, 11: 1–9.

Li, G., Y. Li, G. Han, C. Jiang, M. Geng, N. Guo, W. Wu, S. Liu, Z. Xing, X. Han, and Q. Li. 2024. ’Forecasting and analyzing influenza activity in Hebei Province, China, using a CNN-LSTM hybrid model’, BMC Public Health, 24: 2171.

Li, Q., Z. Zhang, and Z. Ma. 2023. ’Raman spectral pattern recognition of breast cancer: A machine learning strategy based on feature fusion and adaptive hyperparameter optimization’, Heliyon, 9: e18148.

Li, Y., R. M. Reeves, X. Wang, Q. Bassat, W. A. Brooks, C. Cohen, D. P. Moore, M. Nunes, B. Rath, H. Campbell, and H. Nair. 2019. ’Global patterns in monthly activity of influenza virus, respiratory syncytial virus, parainfluenza virus, and metapneumovirus: a systematic analysis’, Lancet Glob Health, 7: e1031–e45.

Liao, C. M., C. F. Chang, and H. M. Liang. 2005. ’A probabilistic transmission dynamic model to assess indoor airborne infection risks’, Risk Anal, 25: 1097–107.

Liu, J., E. Chen, Q. Zhang, P. Shi, Y. Gao, Y. Chen, W. Liu, Y. Qin, Y. Shen, and C. Shi. 2020. ’The correlation between atmospheric visibility and influenza in Wuxi city, China’, Medicine (Baltimore), 99: e21469.

Liu, X., C. Liu, R. Huang, H. Zhu, Q. Liu, S. Mitra, and Y. Wang. 2021. ’Long short-term memory recurrent neural network for pharmacokinetic-pharmacodynamic modeling’, Int J Clin Pharmacol Ther, 59: 138–46.

Lofgren, E., N. H. Fefferman, Y. N. Naumov, J. Gorski, and E. N. Naumova. 2007. ’Influenza seasonality: underlying causes and modeling theories’, J Virol, 81: 5429–36.

Lowen, A. C., and J. Steel. 2014. ’Roles of humidity and temperature in shaping influenza seasonality’, J Virol, 88: 7692–5.

Marr, L. C., J. W. Tang, J. Van Mullekom, and S. S. Lakdawala. 2019. ’Mechanistic insights into the effect of humidity on airborne influenza virus survival, transmission and incidence’, J R Soc Interface, 16: 20180298.

Moriyama, M., W. J. Hugentobler, and A. Iwasaki. 2020. ’Seasonality of Respiratory Viral Infections’, Annu Rev Virol, 7: 83–101.

Moriyama, M., and T. Ichinohe. 2019. ’High ambient temperature dampens adaptive immune responses to influenza A virus infection’, Proc Natl Acad Sci U S A, 116: 3118–25.

Ng, H., Y. Li, T. Zhang, Y. Lu, C. Wong, J. Ni, and Q. Zhao. 2022. ’Association between multiple meteorological variables and seasonal influenza A and B virus transmission in Macau’, Heliyon, 8: e11820.

Park, J. E., W. S. Son, Y. Ryu, S. B. Choi, O. Kwon, and I. Ahn. 2020. ’Effects of temperature, humidity, and diurnal temperature range on influenza incidence in a temperate region’, Influenza Other Respir Viruses, 14: 11–18.

Peci, A., A. L. Winter, Y. Li, S. Gnaneshan, J. Liu, S. Mubareka, and J. B. Gubbay. 2019. ’Effects of Absolute Humidity, Relative Humidity, Temperature, and Wind Speed on Influenza Activity in Toronto, Ontario, Canada’, Appl Environ Microbiol, 85.

Polozov, I. V., L. Bezrukov, K. Gawrisch, and J. Zimmerberg. 2008. ’Progressive ordering with decreasing temperature of the phospholipids of influenza virus’, Nat Chem Biol, 4: 248–55.

Ryu, S., and B. J. Cowling. 2021. ’Human Influenza Epidemiology’, Cold Spring Harb Perspect Med, 11.

Sagripanti, J. L., and C. D. Lytle. 2007. ’Inactivation of influenza virus by solar radiation’, Photochem Photobiol, 83: 1278–82.

Shoji, M., K. Katayama, and K. Sano. 2011. ’Absolute humidity as a deterministic factor affecting seasonal influenza epidemics in Japan’, Tohoku J Exp Med, 224: 251–6.

Si, Xiaohan, Kerrie Mengersen, Chuchu Ye, and Wenbiao Hu. 2024. ’Interactive effect of air pollutant and meteorological factors on seasonal influenza transmission, Shanghai, China’, Atmospheric Environment, 318: 120208.

Soebiyanto, R. P., F. Adimi, and R. K. Kiang. 2010. ’Modeling and predicting seasonal influenza transmission in warm regions using climatological parameters’, PLoS One, 5: e9450.

Soebiyanto, R. P., D. Gross, P. Jorgensen, S. Buda, M. Bromberg, Z. Kaufman, K. Prosenc, M. Socan, T. Vega Alonso, M. A. Widdowson, and R. K. Kiang. 2015. ’Associations between Meteorological Parameters and Influenza Activity in Berlin (Germany), Ljubljana (Slovenia), Castile and León (Spain) and Israeli Districts’, PLoS One, 10: e0134701.

Sooryanarain, H., and S. Elankumaran. 2015. ’Environmental role in influenza virus outbreaks’, Annu Rev Anim Biosci, 3: 347–73.

Tamerius, J. D., J. Shaman, W. J. Alonso, K. Bloom-Feshbach, C. K. Uejio, A. Comrie, and C. Viboud. 2013. ’Environmental predictors of seasonal influenza epidemics across temperate and tropical climates’, PLoS Pathog, 9: e1003194.

Urashima, M., T. Segawa, M. Okazaki, M. Kurihara, Y. Wada, and H. Ida. 2010. ’Randomized trial of vitamin D supplementation to prevent seasonal influenza A in schoolchildren’, Am J Clin Nutr, 91: 1255–60.

Uyeki, T. M., D. S. Hui, M. Zambon, D. E. Wentworth, and A. S. Monto. 2022. ’Influenza’, Lancet, 400: 693–706.

Venna, Rama Krishna Reddy, Amirhossein Tavanaei, Raju N. Gottumukkala, Vijay V. Raghavan, and Stephen Nichols. 2018. ’A Novel Data-driven Model for Real-Time Influenza Forecasting’, IEEE Access.

Wang, C., Y. Li, W. Feng, K. Liu, S. Zhang, F. Hu, S. Jiao, X. Lao, H. Ni, and G. Xu. 2017. ’Epidemiological Features and Forecast Model Analysis for the Morbidity of Influenza in Ningbo, China, 2006-2014’, Int J Environ Res Public Health, 14.

Wang, D., H. Lei, D. Wang, Y. Shu, and S. Xiao. 2023. ’Association between Temperature and Influenza Activity across Different Regions of China during 2010-2017’, Viruses, 15.

Xiao, J., M. Gao, M. Huang, W. Zhang, Z. Du, T. Liu, X. Meng, W. Ma, and S. Lin. 2022. ’How do El Niño Southern Oscillation (ENSO) and local meteorological factors affect the incidence of seasonal influenza in New York state’, Hyg Environ Health Adv, 4.

Xu, D. L., X. K. Hu, and Y. F. Tian. 2017. ’Effect of temperature and food restriction on immune function in striped hamsters (Cricetulus barabensis)’, J Exp Biol, 220: 2187–95.

Yang, W., S. Elankumaran, and L. C. Marr. 2012. ’Relationship between humidity and influenza A viability in droplets and implications for influenza’s seasonality’, PLoS One, 7: e46789.

Zhang, R., Z. Peng, Y. Meng, H. Song, S. Wang, P. Bi, D. Li, X. Zhao, X. Yao, and Y. Li. 2022. ’Temperature and influenza transmission: Risk assessment and attributable burden estimation among 30 cities in China’, Environ Res, 215: 114343.

Zhang, Y., C. Ye, J. Yu, W. Zhu, Y. Wang, Z. Li, Z. Xu, J. Cheng, N. Wang, L. Hao, and W. Hu. 2020. ’The complex associations of climate variability with seasonal influenza A and B virus transmission in subtropical Shanghai, China’, Sci Total Environ, 701: 134607.

Zhao, Z., M. Zhai, G. Li, X. Gao, W. Song, X. Wang, H. Ren, Y. Cui, Y. Qiao, J. Ren, L. Chen, and L. Qiu. 2023. ’Study on the prediction effect of a combined model of SARIMA and LSTM based on SSA for influenza in Shanxi Province, China’, BMC Infect Dis, 23: 71.

Zhou, L., H. Yang, W. Pan, J. Xu, Y. Feng, W. Zhang, Z. Shao, T. Li, S. Li, T. Huang, C. Wang, W. Li, M. Li, S. He, Y. Zhan, and M. Pan. 2022. ’Association between meteorological factors and the epidemics of influenza (sub)types in a subtropical basin of Southwest China’, Epidemics, 41: 100650.

Zhu, H., S. Chen, W. Lu, K. Chen, Y. Feng, Z. Xie, Z. Zhang, L. Li, J. Ou, and G. Chen. 2022. ’Study on the influence of meteorological factors on influenza in different regions and predictions based on an LSTM algorithm’, BMC Public Health, 22: 2335.

