## Supplemental Information for "Meteorological Drivers of Influenza A and B Positivity in a Subtropical Chinese City: A Six-Year Surveillance Study Integrating Distributed Lag Non-Linear Models and Deep Learning"

**Supplementary information**

Supplementary Text

The Supplementary Information document includes six additional files. The first additional file outlines the methodology for nucleic acid extraction and real-time RT-PCR protocols, and the specific principles and formulas for the cell structure of the LSTM network, accompanied by a schematic representation in Figure S1 illustrating the LSTM cell architecture. The second additional file includes supplementary result Tables S2 and S3, detailing the descriptive statistics of the meteorological factors analyzed and the quasi- Akaike information criterion (QAIC) results for various degrees of freedom combinations in the DLNM construction. The third additional file elaborates on the linear association analysis conducted in this study, including a description of the methodology and the calculation of Pearson correlation coefficients between various meteorological factors and influenza cases. The fourth additional file details the methods and results of the targeted sensitivity analysis for the DLNM, demonstrating the robustness and stability of the meteorological exposure-lag-response relationships independent of weekly testing volume fluctuations. The fifth additional file details the methods and results describing the construction and implementation of an Autoregressive Integrated Moving Average (ARIMA) model for comparative analysis. The sixth additional file provides a detailed account of constructing the eXtreme Gradient Boosting (XGBoost) model as the baseline methodological framework employed in this study for benchmarking LSTM performance in the machine learning domain, including the results obtained and their analysis.

**Additional file 1: Supplementary methods**

**Nucleic acid extraction**

A commercial magnetic bead-based kit (Da’an Gene, Guangzhou, China) was used, following the manufacturer’s protocol. The extracted nucleic acid was dissolved in 50 µL of elution buffer, with concentration and purity measured using a Nanodrop 2000 spectrophotometer (Thermo-Fisher Scientific, Waltham, MA). Acceptable nucleic acid purity ratios were defined as an optical density (OD)260/280 ratio between 1.7 and 2.5, and an OD260/230 ratio between 0.5 and 2.5. The integrity of the extracted nucleic acids was confirmed using an Agilent 2100 Bioanalyzer (Agilent Technologies, Santa Clara, CA), with RNA Integrity Number (RIN) values ≥8 and 28S/18S ratios ≥1.5 considered indicative of high-quality samples.

**Real-time reverse transcription polymerase chain reaction (RT-PCR)**

Qualified nucleic acids underwent one-step real-time fluorescence RT-PCR detection within two hours, utilizing commercially available detection kits for influenza A (H1N1) virus (2009) RNA, seasonal influenza virus H3N2 subtype, and influenza B (Victoria and Yamagata lineages), all purchased from Da’an Gene (Guangzhou, China). The detection procedure was performed on a QuantStudio real-time fluorescence quantitative PCR instrument (Thermo-Fisher Scientific, Waltham, MA), following the manufacturer’s instructions. Each detection batch contained negative, positive, and internal control replicates to ensure the reliability of the results. The PCR results were interpreted adhering to the assay manufacturer’s protocols, with amplification curves and cycle threshold (Ct) values used to determine positivity. Positive results were determined if typical amplification curves were observed in both the internal reference fluorescence channel and the target gene fluorescence channel, with corresponding Ct values within the reference range. Specific positive criteria were as follows: for influenza A H1N1, FAM channel amplification with Ct value ≤38; for H3N2, FAM channel amplification with Ct value ≤38; for influenza B Victoria, FAM channel amplification with Ct value ≤38; for Yamagata, VIC channel amplification with Ct value ≤38.

**Specific principles and formulas for the cell structure of the LSTM network**

The core component of the Long Short-Term Memory (LSTM) network is the LSTM cell. The LSTM network incorporates multiple LSTM cells per layer. Each LSTM cell encompasses three gating mechanisms: the forget gate, the input gate, and the output gate (shown in Supplemental Figure S1). These gating mechanisms collaboratively control the flow and updating of information between different time steps, enabling the LSTM to selectively retain or discard information, thereby effectively processing long sequence data.

The forget gate controls the amount of information retained from the previous time step and discards irrelevant data. Mathematically, it is represented as:

$$f_{t}=\sigma\left( W_{f}\cdot\left[ h_{t-1}, {input}_{t} \right]+b_{f} \right)$$

The input gate determines which information from the current time step is added to the cell state, where $C_{t}^{'}$represents the candidate state that could be incorporated. This gate is governed by:

$i_{t}=\sigma\left( W_{i}\cdot\left[ h_{t-1}, {input}_{t} \right]+b_{i} \right)$ and $C_{t}^{'}=function\left( W_{C}\cdot\left[ h_{t-1}, {input}_{t} \right]+b_{C} \right)$, and the cell state is updated as: $C_{t}=f_{t}\times C_{t-1}^{'}+i_{t}\times C_{t}^{'}$

The output gate modulates the final cell state and controls the transmission of information to the next time step, expressed as:

$O_{t}=\sigma\left( W_{O}\cdot\left[ h_{t-1}, {input}_{t} \right]+b_{O} \right)$ and $h_{t}=O_{t}\cdot tan\left( C_{t} \right)$

where ${input}_{t}$ denotes the input sequence at time $t$, the symbol ∙ represents element-wise multiplication, $h_{t}$ and $h_{t-1}$ are the hidden states at time $t$ and $t-1$ respectively, $C_{t}$ is the cell state at time $t$, $W$ represents weight matrices, and $b$ represents bias vectors. $function$ represents the activation function for the candidate state, which is selected based on hyperparameter tuning.

This structure allows the model to capture long-term dependencies within time-series data while efficiently filtering out irrelevant information through the three gates.

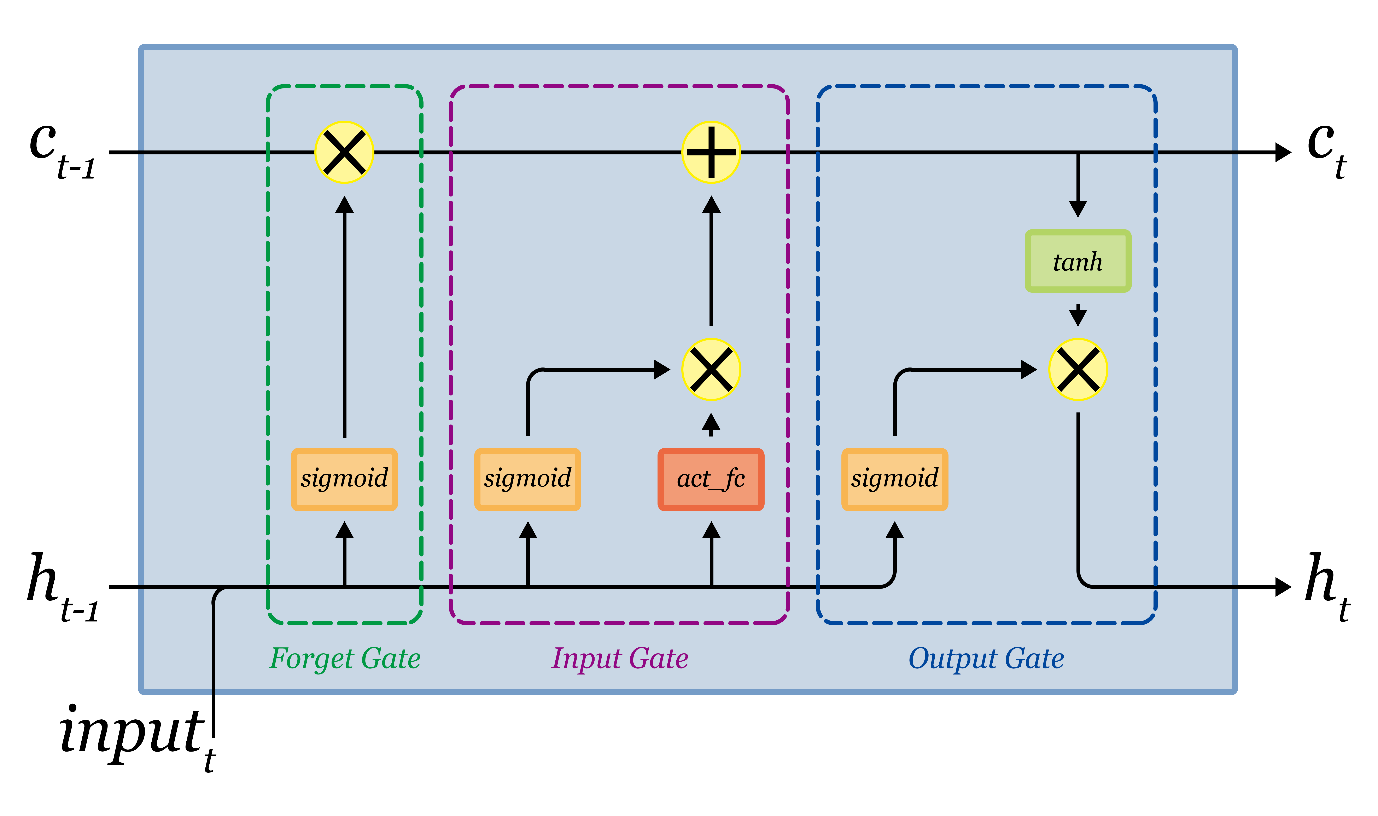

**Figure S1. Detailed Structure of the Long Short-Term Memory (LSTM) Cell.**

**Hyperparameter Optimization Strategy and Search Space**

To ensure optimal convergence and predictive stability of the LSTM networks, we conducted rigorous hyperparameter tuning using the Hyperopt framework. Hyperopt employs a Bayesian optimization strategy based on Tree-based Parzen Estimators (TPE), which significantly outperforms traditional grid search and random search methods by systematically evaluating past performance to propose the next optimal set of hyperparameters (Hanifi et al. 2022). Previous comparative analyses have demonstrated Hyperopt’s superior performance in LSTM applications compared to other optimization libraries, such as Scikit-opt and Optuna (Hanifi, Cammarono, and Zare-Behtash 2024). In our framework, key hyperparameters, including the learning rate, number of hidden layers, number of neurons, and activation functions, were meticulously tuned within established theoretical ranges using Mean Squared Error (MSE) as the reference metric.

**Learning Rate:** Following Bengio, who identified the learning rate as a critical hyperparameter and recommended a search range of 10^-6^ to 1 (Bengio 2012), and corroborated by recent findings demonstrating that rates between 0.0001 and 0.01 yield superior convergence in LSTM networks (Du et al. 2023), we established a discrete learning rate search space of {0.000001, 0.00001, 0.0001, 0.001, 0.01, 0.1}. This specific range balances the trade-off between computational cost and fine-grained optimization.

**Network Architecture:** Theoretically, increasing the number of hidden layers and neurons enhances the algorithm’s fitting capacity; however, excessive complexity risks overfitting and impedes convergence. Based on Panchal’s findings, which suggest that 1 to 2 hidden layers suffice for most applications and 3 layers are considered optimal for maximizing accuracy without severe overfitting (Gaurang et al. 2011), we restricted our architectural exploration to configurations with 1 to 3 hidden layers. Within each hidden layer, the candidate set for the number of neurons was selected as powers of 2 (i.e., {8, 16, 32, 64, 128, 256, 512}) to optimize computational memory efficiency. Furthermore, the batch size for each training session was also tuned covering the range {8, 16, 32, 64, 128, 256, 512}, with the number of epochs flexibly controlled within a natural number interval from 300 to 1000.

**Activation Functions:** Recognizing that no single activation function universally optimizes network performance across all sequential tasks (Jagtap and Karniadakis 2022), we evaluated a comprehensive suite of activation functions. The search space included traditional functions (Sigmoid, Rectified Linear Unit [ReLU], and Hyperbolic Tangent [TanH]) alongside newer, advanced variants (Softplus, Exponential Linear Unit [ELU], and Swish) for automatic selection during the Bayesian optimization process.

**Additional file 2: Supplementary results**

**Table S1:** **Descriptive Statistics of Meteorological Factors in Putian City (2018-2023).**

| Meteorological factor | minimal | 5^th^ percentile | Median | Mean | standard deviation | 95^th^ percentile | maximum |
| --- | --- | --- | --- | --- | --- | --- | --- |
| Average temperature (°C) | 7.6 | 13.8 | 24.1 | 23.3 | 5.6 | 30.7 | 32.9 |
| Humidity (%) | 31 | 57.95 | 75.8 | 76.1 | 10.8 | 94.15 | 99.8 |
| Precipitation (mm) | 0 | 0 | 0 | 3.9 | 10.9 | 23.394 | 144.41 |
| Wind speed (km/h) | 6.8 | 10.8 | 18 | 17.9 | 5.4 | 25.2 | 116.8 |
| Wind direction * | NW | - | SW | - | - | - | E |
| Solar radiation (W/m^2^) | 5.8 | 73.1 | 205.3 | 202.0 | 69.1 | 305.9 | 341.3 |
| Ultraviolet index | 0 | 3 | 7 | 7.1 | 2.1 | 10 | 10 |
| Daily temperature ranges (°C) | 1 | 3 | 8 | 7.8 | 3.0 | 13 | 16 |

*Note: Wind direction is described based on the frequency of occurrence of the eight cardinal directions. NW: north-westerly; SW: south-westerly; E: easterly.

**Table S2: QAIC Values for DLNM Models Across Degrees of Freedom Combinations.**

| Influenza type | Meteorological factor | ${df}_{x}$ | $\mathrm{df}_{\mathrm{time}}$ | | | | |
| --- | --- | --- | --- | --- | --- | --- | --- |
|  |  |  | 6 | 7 | 8 | 9 | 10 |
| influenza A | humidity | 2 | - | 1.04E+36 | 7921.38 | 4113.21 | 3170.75 |
|  |  | 3 | 12562353 | 3317.23 | 10240.29 | 5143.05 | 3153.05 |
|  |  | 4 | 26540151 | 9.74E+17 | 42517.57 | 81578405 | 3096.05 |
|  |  | 5 | 3.11E+08 | 3253.86 | 494344.9 | 3442.48 | 3068.09 |
|  | precipitation | 2 | - | - | - | - | - |
|  |  | 3 | 51466.64 | 3531.96 | 4224.61 | 3933.90 | 3103.71 |
|  |  | 4 | 4.19E+134 | 3535.98 | 4834.92 | 4409.85 | 3103.31 |
|  |  | 5 | 210749.33 | 3572.73 | 5381.03 | 4498.39 | 3104.24 |
|  | solar radiation | 2 | 213743.68 | 3255.26 | 4605.74 | 3623.64 | 3181.35 |
|  |  | 3 | - | 2.1E+120 | 5183.57 | 3792.24 | 3181.52 |
|  |  | 4 | - | 3246.15 | 4806.17 | 3753.06 | 3173.27 |
|  |  | 5 | 239783 | 3243.05 | 6172.39 | 3773.43 | 3167.59 |
|  | daily temperature range | 2 | - | 3340.64 | 4410.01 | 3497.04 | 3169.77 |
|  |  | 3 | 293680.27 | 3384.87 | 4808.02 | 3349.16 | 3175.01 |
|  |  | 4 | - | 1.07E+173 | 4833.78 | 3325.88 | 3160.23 |
|  |  | 5 | - | 3384.08 | 5610.18 | 3297.89 | 3123.95 |
|  | temperature | 2 | 40091169 | 3313.98 | 4954.56 | 1.58E+40 | 3188.91 |
|  |  | 3 | 303643.15 | 3218.40 | 4898.15 | 4195.35 | 3096.06 |
|  |  | 4 | 68152.83 | 3240.96 | 4327.51 | 3769.00 | 3104.44 |
|  |  | 5 | - | 3252.09 | 4135.43 | 1.59E+256 | 3089.88 |
|  | ultraviolet index | 2 | 211637.62 | 3207.10 | 4757.41 | 32337.63 | 3156.51 |
|  |  | 3 | 152137.84 | 3194.74 | 4386.31 | 3540.17 | 3154.23 |
|  |  | 4 | 1.7E+09 | 3197.19 | 5059.73 | 4097.16 | 3136.33 |
|  |  | 5 | 61743113 | 3194.97 | 3217.16 | 3225.33 | 3122.32 |
|  | wind direction | 2 | 215264.74 | 5.2E+118 | 4243.30 | 3730.69 | 3135.29 |
|  |  | 3 | - | 7.81E+185 | 4232.54 | 7.96E+08 | 3105.84 |
|  |  | 4 | 4.16E+16 | 3249.18 | 3817.01 | 7.07E+91 | 3097.46 |
|  |  | 5 | 2.1E+109 | 3211.63 | 3842.97 | 3324.891 | 3093.36 |
|  | wind speed | 2 | 370034.51 | 3344.84 | 4504.67 | 2.33E+20 | 3177.68 |
|  |  | 3 | 409397.77 | 3447.91 | 4404.46 | 4491.16 | 3134.36 |
|  |  | 4 | 4.75E+16 | 3494.94 | 4729.04 | 5548.65 | 3129.81 |
|  |  | 5 | 539380.65 | 539306.70 | 4193.76 | 4.17E+18 | 3122.94 |
| influenza B | humidity | 2 | 3171.40 | - | 3175.68 | 3173.96 | - |
|  |  | 3 | 3175.22 | - | 3176.62 | 3173.47 | 3232.91 |
|  |  | 4 | 3115.23 | - | 3120.42 | 3117.40 | 3095.73 |
|  |  | 5 | 3077.17 | - | 2.91E+184 | 3081.48 | 3060.79 |
|  | precipitation | 2 | - | - | - | - | - |
|  |  | 3 | 3119.93 | - | 3102.16 | 3097.36 | 3072.71 |
|  |  | 4 | 3059.24 | - | 3012.14 | 3003.45 | 2983.99 |
|  |  | 5 | 3054.20 | - | 2993.22 | 2992.69 | 2963.95 |
|  | solar radiation | 2 | 3062.49 | - | 3058.97 | 3049.41 | 3028.98 |
|  |  | 3 | 3048.52 | - | 3040.77 | 3023.35 | 1.43E+54 |
|  |  | 4 | 3026.96 | - | 3028.70 | 3001.70 | - |
|  |  | 5 | 3029.44 | - | 1.91E+80 | 3005.49 | 3013.18 |
|  | daily temperature range | 2 | 3112.52 | - | 3116.68 | 3121.76 | 1.21E+138 |
|  |  | 3 | 3100.92 | 4.06E+12 | 3101.86 | 3101.91 | 7.24E+09 |
|  |  | 4 | 3018.37 | - | 3.12E+13 | 3013.11 | 3008.44 |
|  |  | 5 | 2985.16 | 6.89E+262 | 9.54E+221 | 2977.85 | 2973.01 |
|  | temperature | 2 | 3104.50 | - | - | 3049.23 | 3038.55 |
|  |  | 3 | 3048.00 | - | - | 3022.72 | 6.91E+84 |
|  |  | 4 | 3040.63 | - | 3047.21 | 3010.52 | 2997.21 |
|  |  | 5 | 3039.46 | 5.26E+152 | 3045.00 | 3008.85 | 2.5E+108 |
|  | ultraviolet index | 2 | 3084.96 | - | 3082.84 | 3066.21 | 3033.36 |
|  |  | 3 | 3101.43 | - | 3080.68 | 3060.74 | 1.58E+152 |
|  |  | 4 | 3185.54 | - | 3038.69 | 2992.05 | - |
|  |  | 5 | 3077.97 | - | 2983.66 | 2980.61 | 1.7E+113 |
|  | wind direction | 2 | 3159.66 | 8.1E+27 | 3155.93 | 3099.48 | 5375.50 |
|  |  | 3 | 3160.55 | 4.07E+121 | - | 3071.69 | 3046.36 |
|  |  | 4 | 3213.52 | - | 3101.80 | 3047.93 | 3016.08 |
|  |  | 5 | 3161.81 | 5.50E+229 | 3091.10 | 3042.39 | 2996.10 |
|  | wind speed | 2 | 3199.67 | 21479.97 | 3204.81 | 3190.44 | 3.37E+130 |
|  |  | 3 | 3168.48 | - | - | 3167.41 | 4.2E+99 |
|  |  | 4 | 3152.88 | 3139.90 | 3173.00 | 3159.70 | 3.51E+171 |
|  |  | 5 | 3144.57 | - | 3165.02 | 3155.37 | 2.5E+108 |

Note: DLNM (Distributed Lag Non-linear Models), QAIC (quasi- Akaike information criterion), Degrees of Freedom ($df$). ‘-’ indicates poor model fit or non-convergence at the specified $df$ combination, precluding QAIC calculation.

**Additional file 3: Linear association analysis**

**Method**

Pearson correlation analysis was conducted to assess the linear relationships between meteorological factors and influenza incidence, with correlation coefficient matrices plotted to reveal underlying association patterns.

**Result**

In this study, Pearson correlation coefficients were calculated between various meteorological factors and influenza cases, and scatter plots, density plots and correlation coefficient matrices were generated. The analysis revealed that influenza A cases were statistically significantly negatively correlated with average temperature (r = -0.163), wind speed (r = -0.093), solar radiation (r = -0.100), UV index (r = -0.072), and wind direction (r = -0.064). Similarly, influenza B cases also exhibited comparable negative correlations with these meteorological factors: average temperature (r = -0.197), wind speed (r = -0.091), solar radiation (r = -0.063), UV index (r = -0.071), and wind direction (r = -0.078). These results revealed that the linear correlations between the meteorological factors and the number of influenza A cases, influenza B cases and influenza-like illness cases were weak (as shown in Figure S2).

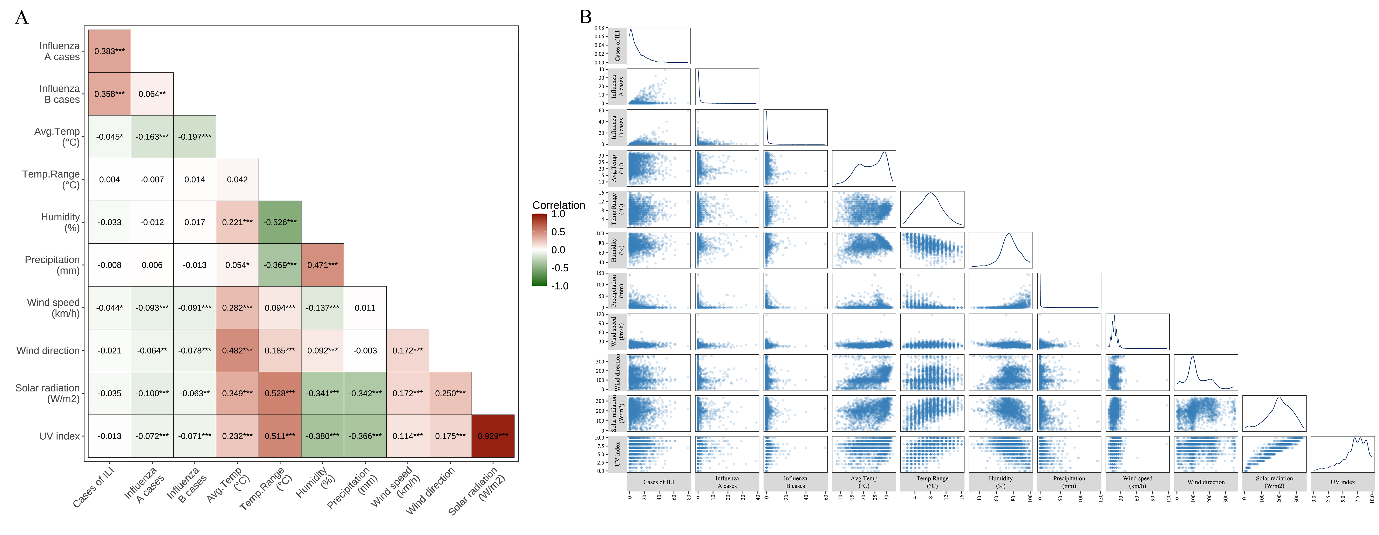

**Figure S2. Correlation Heatmap and Scatterplot Matrix Illustrating Pairwise Associations and Potential Collinearity among Influenza Indicators and Meteorological Variables.** This composite plot visualizes the complex relationships between influenza-like illness (ILI) cases, influenza A and B positive rates, and multiple meteorological variables, including average temperature, humidity, precipitation, wind speed, wind direction, solar radiation, and ultraviolet (UV) index.

**(A) Pearson Correlation Heatmap:** This section presents a correlation coefficient matrix, with color intensity reflecting the strength of correlations between the variables. Red cells represent positive correlations, while green cells represent negative correlations. Darker shades correspond to correlation coefficients approaching 1 or -1, indicating stronger linear relationships, with statistically significant correlations marked by asterisks (*). This matrix allows for a rapid identification of notable relationships and potential collinearity (e.g., between solar radiation and UV index) among input features.

**(B) Scatterplot Matrix and Density Distributions:** (1) Lower-left triangle: This section contains scatterplots of pairwise comparisons between variables. These plots facilitate the visual identification of potential linear or non-linear epidemiological relationships, as well as any outliers or anomalies. This visualization is essential for evaluating the nature of interactions between meteorological factors and influenza cases.

(2) Diagonal: The diagonal displays the density distribution curves for each individual variable. These curves provide an overview of the distribution characteristics of each variable, revealing central tendencies, variance, and any skewness present within the longitudinal data.

**Additional file 4: Sensitivity Analysis of the DLNM Model**

**Rationale and methodological framework**

In the principal DLNM analysis, weekly testing volumes were incorporated as a covariate to account for variations in surveillance intensity that could potentially confound the estimated associations between meteorological factors and influenza positivity rates. To rigorously assess the robustness of the DLNM findings against the influence of this covariate and to determine whether the principal conclusions are sensitive to its inclusion, we conducted a comprehensive sensitivity analysis by reconstructing the DLNM with the weekly detection variable removed while preserving all other model specifications.

**Sensitivity analysis methodology**

The sensitivity analysis was implemented in two complementary dimensions. First, we evaluated the impact of weekly detection volumes variable removal on the 15-day cumulative relative risk estimates across all meteorological factors for both influenza A and B subtypes. Second, we examined the lag-specific relative risk patterns under extreme meteorological conditions (defined as the 95^th^ and 5^th^ percentiles of each variable) following variable removal. The reconstructed DLNMs retained identical degrees of freedom for the cross-basis splines, lag structure (15-day maximum lag), reference values, and remaining covariates as those specified in the original model. Comparisons between the reconstructed and original models focused on the directionality, magnitude, and statistical significance of the estimated effects.

**Results of cumulative risk sensitivity analysis**

The sensitivity analysis for cumulative relative risk estimates demonstrated substantial concordance between the reconstructed DLNM (with weekly detection volumes variable removed) and the original model. The non-linear response patterns of temperature, humidity, precipitation, wind speed, solar radiation, and daily temperature range on the 15-day cumulative risks of both influenza A and B positivity rates remained largely consistent in shape, magnitude, and statistical significance (Figure S3). Two notable deviations were observed: (1) for influenza A, the ultraviolet index exhibited a non-monotonic trajectory in the reconstructed model, initially increasing before declining, whereas the original model showed a continuous decreasing trend; however, the increasing segment in the reconstructed model did not reach statistical significance; (2) for influenza B, the protective effect of wind speeds exceeding 28 km/h on cumulative risk lost statistical significance in the reconstructed model, whereas this protective association was statistically significant in the original model. These minor deviations notwithstanding, the principal findings regarding the dominant meteorological drivers of influenza A (warm-humid conditions) and influenza B (cool-wet conditions) remained robust to the removal of the weekly detection variable.

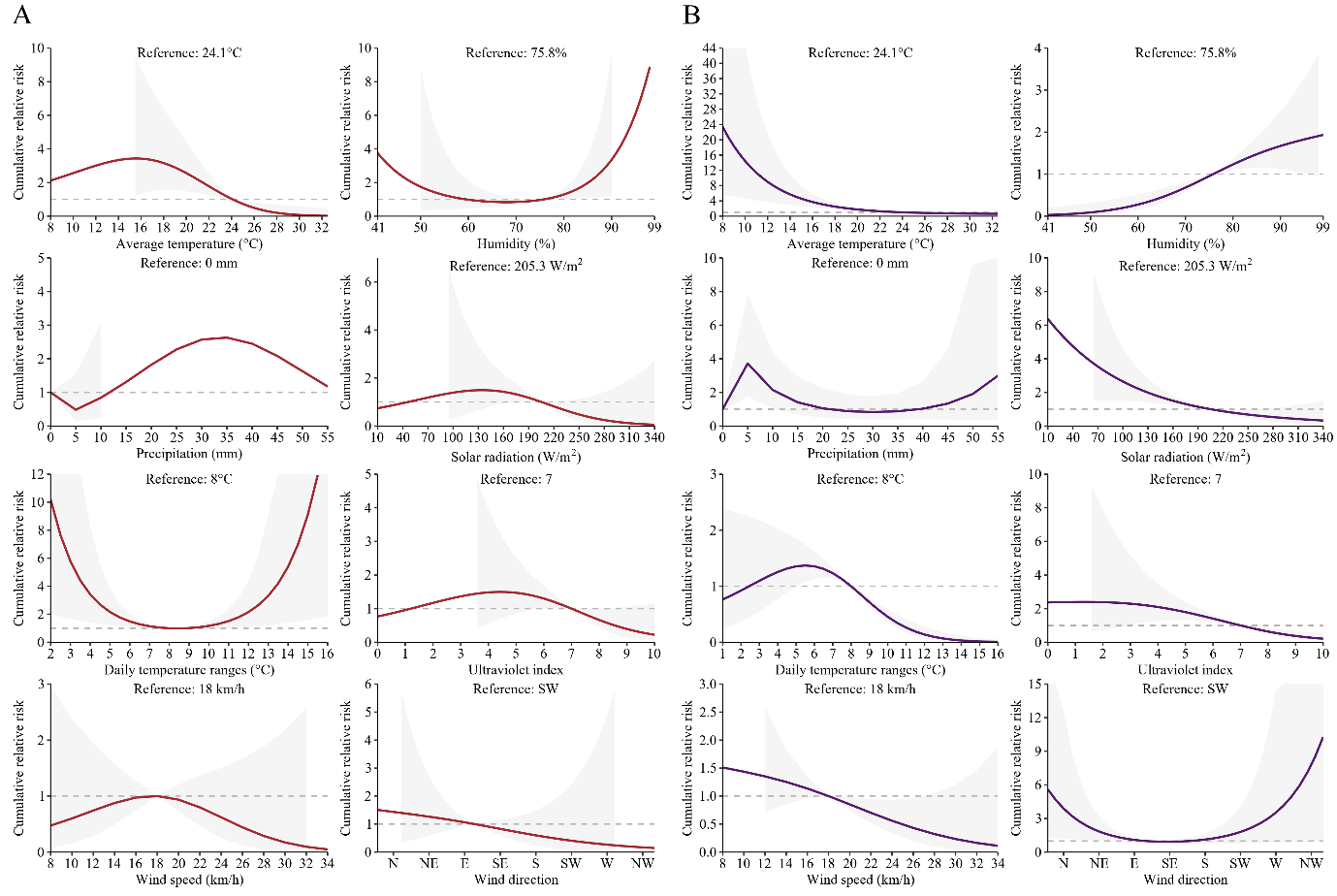

**Figure S3. Sensitivity Analysis of the Cumulative Risk of Meteorological Factors on Influenza A and B Positivity Rates.** This figure illustrates the non-linear, cumulative exposure-response relationships between eight individual meteorological drivers and the 15-day cumulative relative risk of influenza A and B positivity, estimated via a reconstructed Distributed Lag Non-linear Model (DLNM). Unlike the primary analysis (Main Text Figure 4), this sensitivity model intentionally excludes the “weekly detection volumes” covariate to assess the potential impact of surveillance intensity bias. Panels in **(A)** depict the non-linear cumulative risks for influenza A, while panels in **(B)** show the corresponding risks for influenza B. In each subplot, the x-axis represents the observed continuous range of a specific meteorological factor, and the y-axis indicates the cumulative relative risk compared to a predefined median reference value. The solid line reflects the estimated non-linear trend, and shaded regions denote the 95% confidence intervals (CIs). The structural consistency between these curves and those in Figure 4 empirically confirms that the identified meteorological drivers are intrinsic environmental triggers, independent of testing volume fluctuations.

**Results of extreme condition lag-effect sensitivity analysis**

The sensitivity analysis for lag-specific relative risk patterns under extreme meteorological conditions revealed that the reconstructed DLNM produced results substantially consistent with those of the original model (Figure S4). The temporal heterogeneity of meteorological effects on influenza A and B positivity rates, including the delayed peak effects of extreme high temperature on influenza A and the sustained risk elevations under extreme low temperature for influenza B, was preserved across both modeling configurations. The lag-response patterns for extreme precipitation, humidity, wind speed, solar radiation, ultraviolet index, and daily temperature range exposures similarly demonstrated qualitative and quantitative agreement between the original and reconstructed models.

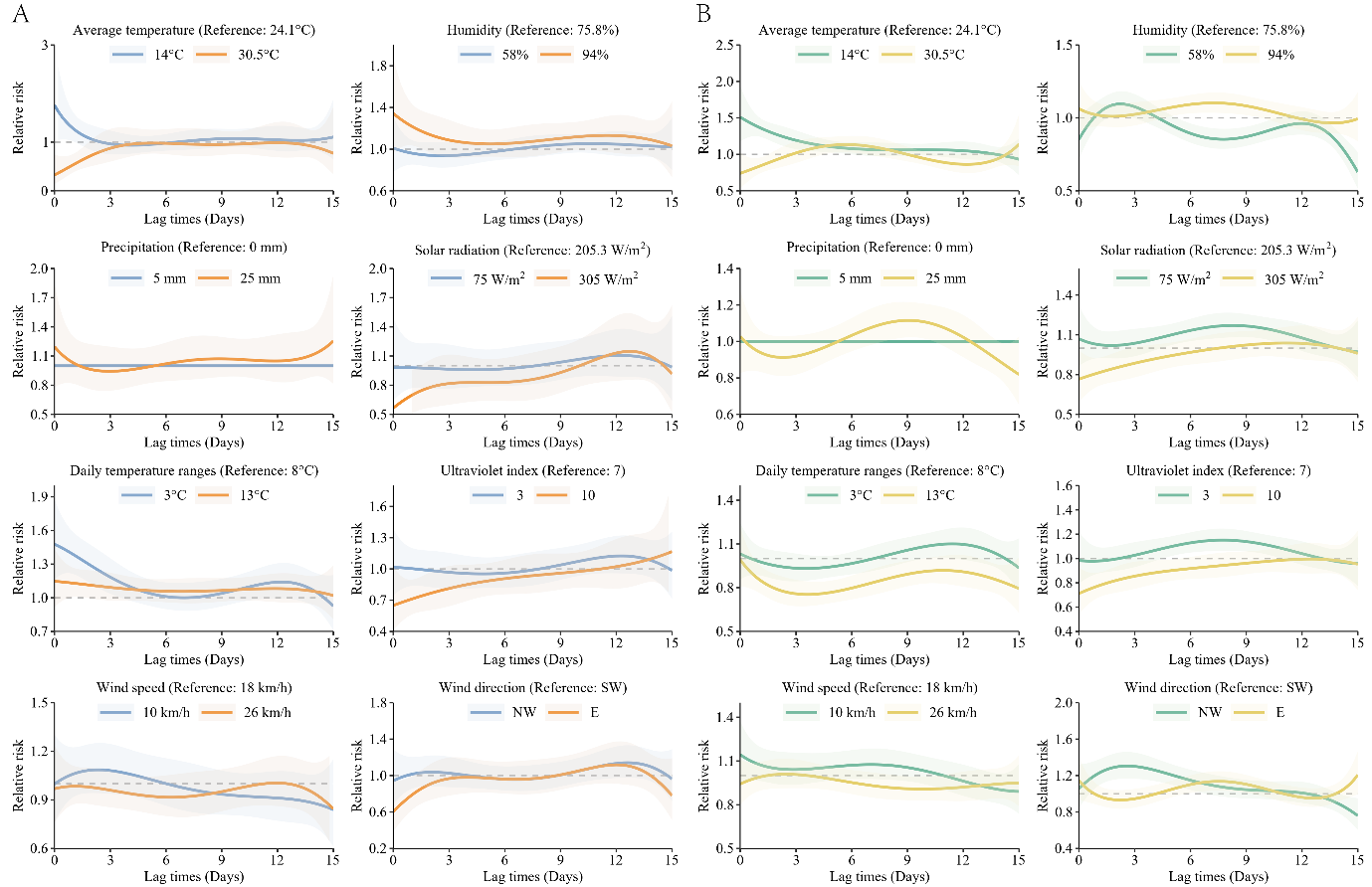

**Figure S4. Sensitivity Analysis of the Lagged Temporal Effects of Extreme Meteorological Conditions on Influenza A and B Positivity Rates.** This figure visualizes the distribution of relative risks over a 15-day lag period following exposure to extreme meteorological conditions, derived from a reconstructed DLNM. Similar to Figure S3, this sensitivity model excludes the “weekly detection volumes” covariate. The analysis compares the temporal lag effects at the extreme high (95^th^ percentile) and extreme low (5^th^ percentile) values of selected weather variables for influenza A **(A)** and influenza B **(B)**. In each subplot, the x-axis represents the specific lag days (from day 0 to 15), and the y-axis shows the relative risk compared to the median reference condition. Solid orange lines (influenza A) and yellow lines (influenza B) represent extreme high-value conditions, whereas solid blue lines (influenza A) and green lines (influenza B) represent extreme low-value conditions, with shaded areas representing 95% CIs. The robust concordance of these lag-response patterns with Main Text Figure 5 further substantiates that extreme weather events exert genuine, time-lagged epidemiological effects that are not artifacts of weather-correlated clinical testing behaviors.

**Interpretation and implications**

Collectively, the sensitivity analysis findings substantiate the robustness of the principal DLNM conclusions regarding type-specific meteorological drivers of influenza transmission. The minor deviations observed in select parameters (ultraviolet index for influenza A; wind speed for influenza B) suggest that while weekly testing volumes may exert subtle modifying effects on certain associations, the overall meteorological response architecture identified in the original analysis is not critically dependent on the inclusion of this covariate. These results enhance confidence in the validity of the meteorological driver characterizations presented in the main analysis and support the interpretive framework adopted in the principal manuscript.

**Additional file 5: The construction and analysis of** **Autoregressive Integrated Moving Average (ARIMA) model**

**Method**

To establish a mathematically fair baseline against the deep learning framework, the conventional Autoregressive Integrated Moving Average (ARIMA) model was extended to a multivariate formulation to incorporate external regressors. The model was fitted using the auto.arima() function from the R package forecast. Crucially, the identical multidimensional covariate matrix used in the LSTM, comprising meteorological factors, weekly detection volumes, mask-wearing stringency indices, day of the week (DOW), and non-local population proportions, was explicitly fed into the model via the xreg argument. The function autonomously optimized the autoregressive, differencing, and moving average parameters (p, d, q) based on the Akaike Information Criterion (AIC/QAIC). Data partitioning was strictly consistent with the LSTM pipeline: 2018–2022 data were utilized for training, while 2023 data served as the validation set for predicting Influenza A and B positivity rates. Performance was assessed using Mean Absolute Error (MAE), Root Mean Square Error (RMSE), Mean Absolute Percentage Error (MAPE), and Symmetric Mean Absolute Percentage Error (SMAPE), and compared with that from the Bayesian-optimized Long Short-Term Memory (LSTM) neural network.

The four metrics, MAE, RMSE, MAPE, and SMAPE are calculated as:

MAE = $\frac{1}{n}\sum_{i=1}^{n} |y_{i}-\hat{y}_{i}|$

RMSE = $\sqrt{\frac{1}{n}\sum_{i=1}^{n} (y_{i}-\hat{y}_{i})^{2}}$

MAPE = $\frac{1}{n}\sum_{i=1}^{n} \left| \frac{y_{i}-\hat{y}_{i}}{y_{i}} \right|\times100$

SMAPE = $\frac{1}{n}\sum_{i=1}^{n} \frac{|y_{i}-\hat{y}_{i}|}{(y_{i}+\hat{y}_{i})/2}\times100$

with adjustments for zero denominators.

**Results**

The MAE for the covariate-adjusted ARIMA model of influenza A was 0.136, RMSE was 0.238, MAPE was 1.115, and SMAPE was 1.212. For influenza B, the MAE was 0.049, RMSE was 0.057, MAPE was 1.426, and SMAPE was 0.810. Overall, the predictive performance of the ARIMA model was substantially inferior to that of the Bayesian-optimized LSTM. As illustrated in Figure S5, the linear model struggled significantly with the non-stationary dynamics of the 2023 viral rebound. It either failed entirely to capture the extreme, explosive non-linear peaks (as seen in Influenza A) or generated continuous spurious predictions during zero-case periods due to mechanical linear reactions to covariate inputs (as seen in Influenza B).

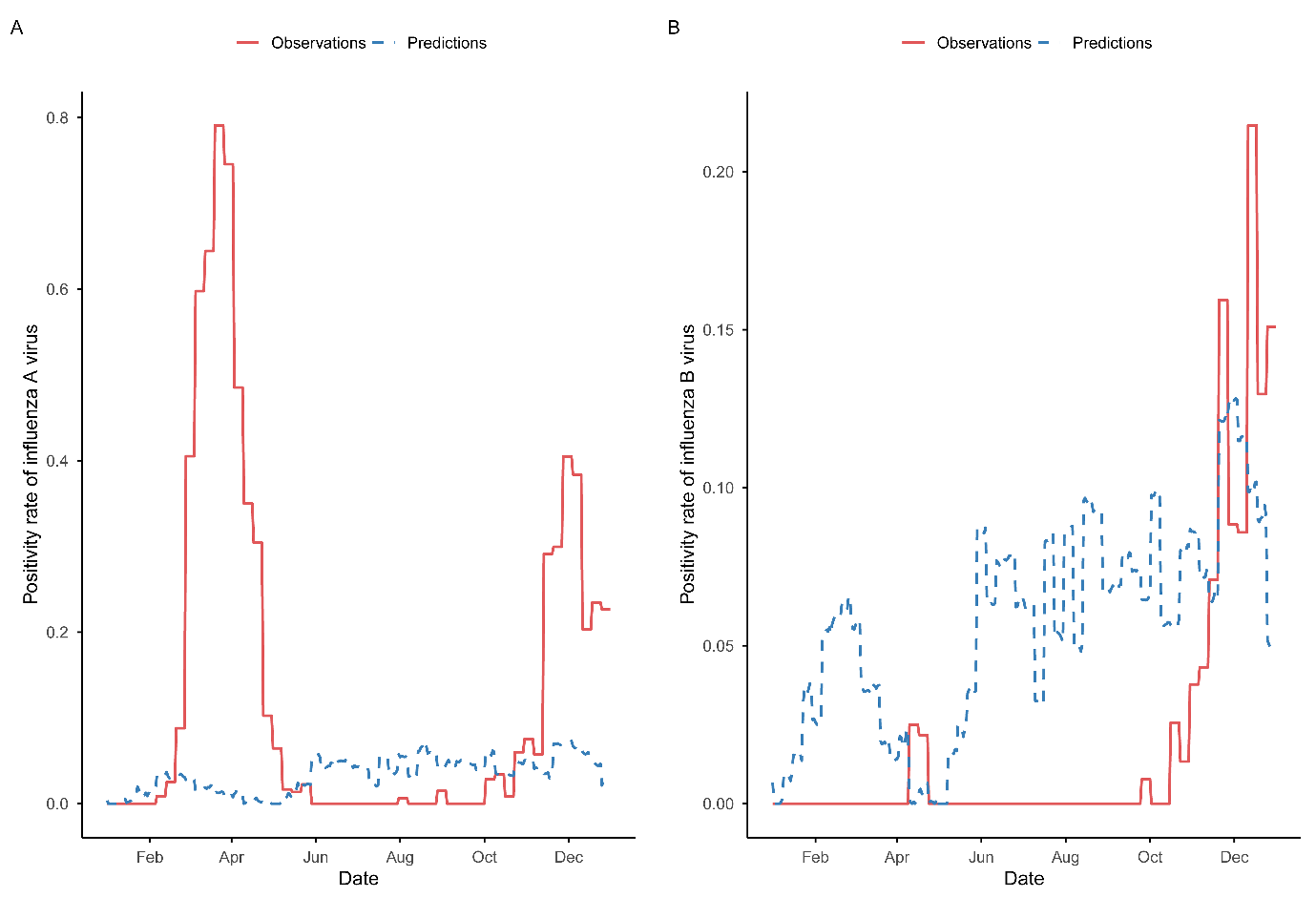

**Figure S5. The Actual Values of Influenza A and B Compared to the Predicted Values from the Covariate-Adjusted ARIMA Model.** The visualizations explicitly reveal the fundamental limitations of applying a linear autoregressive model to non-stationary, multi-covariate pandemic-era data. **(A)** For influenza A, the ARIMA model predicts relatively flat, conservatively smoothed values, failing entirely to capture the extreme, explosive non-linear peaks associated with the 2023 viral rebound. **(B)** For influenza B, the model produces continuous, spurious fluctuations during non-epidemic periods (likely a linear overreaction to variations in exogenous covariates like testing volumes or weather factors), while simultaneously failing to capture the true magnitude of the winter peak. Solid red lines represent actual observed positivity rates, while dashed blue lines indicate predicted values. Note: ARIMA refers to the multivariate Autoregressive Integrated Moving Average model incorporating the exact same exogenous variables (meteorology, NPIs, testing volumes) as the LSTM framework.

**Additional file 6: Comparative Analysis with an eXtreme Gradient Boosting (XGBoost) Machine-Learning Baseline**

**Rationale**

While the multivariate Autoregressive Integrated Moving Average (ARIMA) model detailed in Additional File 5 constitutes the canonical linear statistical benchmark for epidemiological time-series forecasting, gradient-boosted decision-tree ensembles, most notably eXtreme Gradient Boosting (XGBoost), have emerged as one of the leading non-deep-learning approaches for structured predictive tasks (Chen and Guestrin 2016). To subject our Bayesian-optimised Long Short-Term Memory (LSTM) network to a rigorous machine-learning benchmark and to isolate the incremental predictive value contributed by the recurrent architecture itself, we constructed an XGBoost regression model using the identical multidimensional covariate matrix employed in the deep-learning pipeline. Importantly, to guarantee methodological comparability, the XGBoost model was supplied with the identical set of exogenous covariates as the Bayesian-optimised LSTM, meteorological variables, weekly testing volumes, mask-wearing stringency indices, migrant population proportions, and day-of-the-week indicators, without any additional lag features, autoregressive terms, or engineered temporal representations, so as to strictly isolate the predictive contribution attributable to sequential inductive bias.

**Methods**

**Model implementation.** XGBoost provides a scalable gradient-boosting framework built upon sparse classification and regression trees (CART), representing the state-of-the-art non-deep-learning approach for structured predictive tasks. The model was fitted in R (xgboost package, version 3.2.1.1).

**Input feature space.** The predictor set was strictly matched to the LSTM covariate architecture and comprised: (i) eight meteorological variables (average temperature, humidity, precipitation, wind speed, wind direction, solar radiation, ultraviolet index, and daily temperature range); (ii) weekly influenza testing volumes; (iii) the mask-wearing stringency index; (iv) a day-of-week indicator; and (v) the proportion of non-local (migrant) population. Critically, no additional lag features, sliding-window statistics, or autoregressive terms were introduced, thereby ensuring that the information set available to XGBoost was identical to the contemporaneous covariate input consumed by the LSTM at each time step. This design deliberately isolates the predictive contribution of *architectural inductive bias*, specifically, the LSTM’s intrinsic sequential memory versus XGBoost’s memory-free tree-partitioning geometry, from any confounding due to hand-engineered temporal feature construction.

**Data partitioning.** The training-validation split strictly followed the LSTM and ARIMA protocols: data from 2018–2022 constituted the training set, while 2023 served as the held-out validation set for out-of-sample forecasting of weekly Influenza A and B positivity rates.

**Performance evaluation.** Predictive accuracy was quantified using mean absolute error (MAE), root mean squared error (RMSE), mean absolute percentage error (MAPE), and symmetric mean absolute percentage error (SMAPE), calculated identically to the metrics reported for the ARIMA baseline in Additional File 5.

**Results**

The XGBoost model produced substantially higher forecasting errors than the LSTM across both influenza subtypes (Table S3; Figure S6). For **Influenza A**, XGBoost yielded MAE = 0.1379, RMSE = 0.2161, MAPE = 1.8195, and SMAPE = 1.0805, errors of a magnitude broadly comparable to the multivariate ARIMA baseline (MAE = 0.136) and approximately 15-fold higher than the LSTM (MAE = 0.009). For **Influenza B**, XGBoost achieved MAE = 0.0704, RMSE = 0.0739, MAPE = 1.9172, and SMAPE = 0.7371, exceeding both the ARIMA benchmark (MAE = 0.049) and the LSTM (MAE = 0.002) by approximately 35-fold relative to the latter.

**Table S3. Comparative predictive performance of LSTM, multivariate ARIMA, and XGBoost on the 2023 validation set.**

| **Model** | **Subtype** | **MAE** | **RMSE** | **MAPE** | **SMAPE** |
| --- | --- | --- | --- | --- | --- |
| Bayesian-optimised LSTM | Influenza A | 0.009 | 0.035 | 0.158 | 0.521 |
| Multivariate ARIMA | Influenza A | 0.136 | 0.238 | 1.115 | 1.212 |
| XGBoost | Influenza A | 0.138 | 0.216 | 1.820 | 1.081 |
| Bayesian-optimised LSTM | Influenza B | 0.002 | 0.011 | 0.170 | 0.484 |
| Multivariate ARIMA | Influenza B | 0.049 | 0.057 | 1.426 | 0.810 |
| XGBoost | Influenza B | 0.070 | 0.074 | 1.917 | 0.737 |

Note: All three models received the identical multidimensional covariate matrix. Best-performing values are in bold.

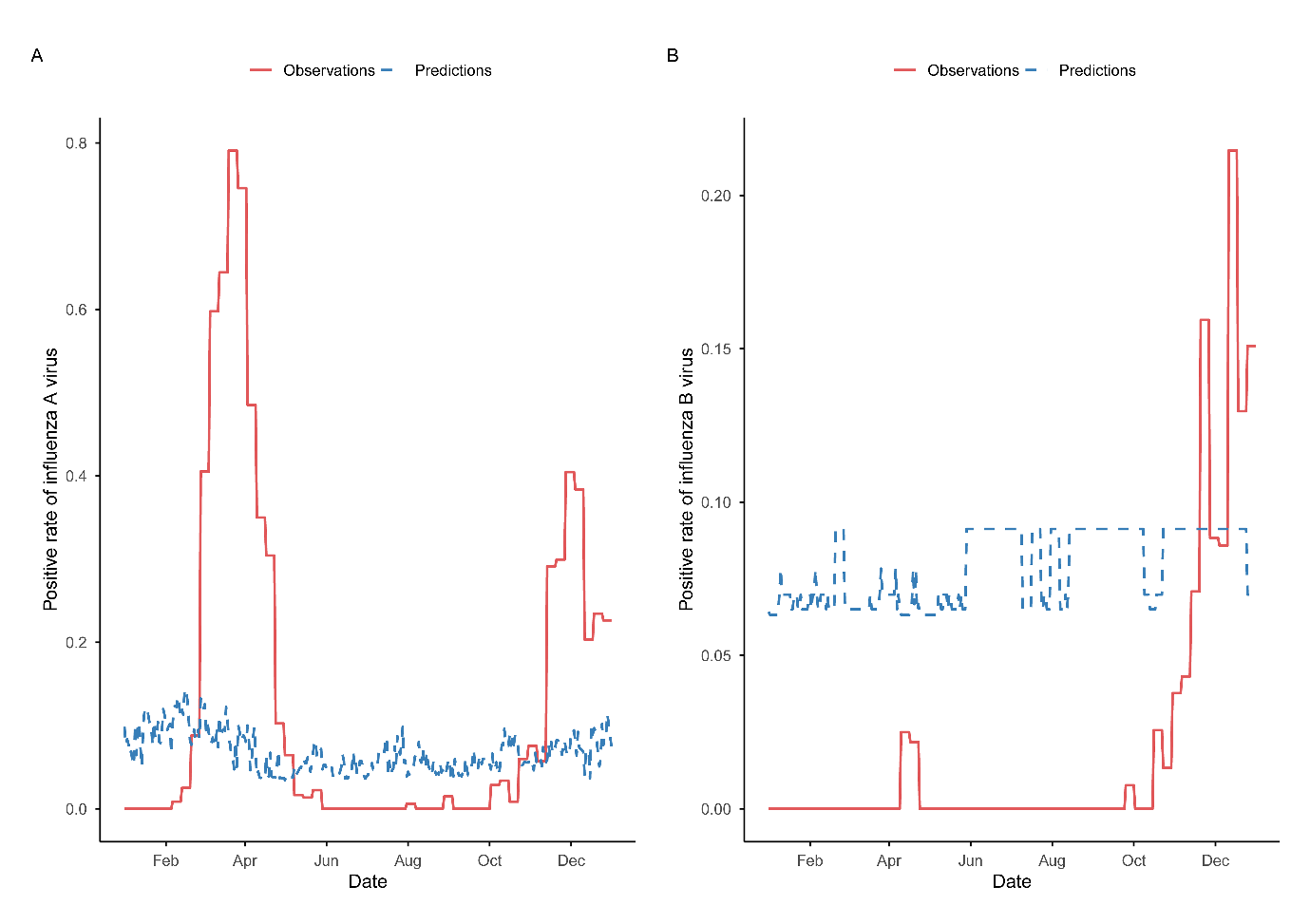

**Figure S6. Observed versus XGBoost-predicted weekly positivity rates for (A) Influenza A and (B) Influenza B in Putian City, 2023.** Solid lines represent observed positivity rates; dashed lines depict XGBoost forecasts. The model systematically under-predicts the explosive Influenza A rebound in February-June and October–December 2023 peaks and produces largely flat forecasts for Influenza B, failing to reproduce the pronounced mid-year 2023 peak that characterised the post-NPI resurgence of type B circulation in the study population.

**Interpretation**

The systematically inferior performance of XGBoost, despite receiving the identical covariate matrix as the LSTM, provides architecturally informative insight into the architectural properties required for post-pandemic influenza forecasting. Four fundamental limitations of the gradient-boosted tree framework, articulated in the Discussion section of the main manuscript, jointly account for this performance gap:

1. **Absence of intrinsic temporal ordering.** Decision trees construct predictions through threshold-based partitions along feature dimensions; their splitting logic is insensitive to the sequential ordering of observations and cannot structurally express the temporal dependency “the current time step depends on prior history” in the manner naturally accommodated by recurrent neural architectures.
2. **Inability to model long-range temporal dependencies.** XGBoost lacks any mechanism analogous to the LSTM’s gated recurrence, through which historical states are propagated, filtered, and dynamically re-weighted according to their relevance to the current prediction. Consequently, XGBoost cannot represent the non-linear, long-horizon temporal dependencies that characterise influenza dynamics.
3. **Violation of the independence assumption.** Tree-based ensembles implicitly assume that samples are independent and identically distributed, an assumption fundamentally at odds with the strong serial autocorrelation intrinsic to epidemiological time series. XGBoost typically neglects the temporal structure of residuals and cannot effectively model serial dependence.
4. **Non-comparable time-modelling paradigms.** The central methodological question of our forecasting analysis was whether a sequential deep-learning model offers substantive gains over traditional approaches. Because XGBoost belongs to a fundamentally non-sequential modelling paradigm, its comparison with the LSTM primarily illuminates the limitations of memory-free machine learning under non-stationary epidemiological conditions, rather than an equivalent “horse race” between competing sequential frameworks. The ARIMA benchmark, which likes the LSTM, is a sequential model that builds temporal dependence directly into its structure, therefore provides the more paradigmatically comparable reference against which the deep-learning architecture’s advantages can be most directly discerned.

These results reinforce our central methodological conclusion: the LSTM’s decisive predictive advantage during the volatile 2023 post-NPI transition arises not from privileged access to information, but from its architectural capacity to represent evolving, non-stationary, autocorrelated dynamics, capacities absent in both linear autoregressive (ARIMA) and non-linear memory-free machine-learning (XGBoost) baselines. Our finding has methodological implications beyond the present dataset: it suggests that recurrent gated architectures constitute a principled design choice for public-health forecasting under regimes of pronounced non-stationarity, such as those induced by the pandemic-era disruption of respiratory-virus circulation.

REFERENCES

Bengio, Yoshua. 2012. "Practical Recommendations for Gradient-Based Training of Deep Architectures." In *Neural Networks*.

Chen, Tianqi, and Carlos Guestrin. 2016. "XGBoost: A Scalable Tree Boosting System." In *Proceedings of the 22nd ACM SIGKDD International Conference on Knowledge Discovery and Data Mining*, 785–94. San Francisco, California, USA: Association for Computing Machinery.

Du, X., L. Si, P. Li, and Z. Yun. 2023. 'A method for detecting the quality of cotton seeds based on an improved ResNet50 model', *PLoS One*, 18: e0273057.

Gaurang, Panchal, Amit Ganatra, Y. Kosta, and Devyani Panchal. 2011. 'Behaviour Analysis of Multilayer Perceptronswith Multiple Hidden Neurons and Hidden Layers', *International Journal of Computer Theory and Engineering*, 3: 332-37.

Hanifi, Shahram, Andrea Cammarono, and Hossein Zare-Behtash. 2024. 'Advanced hyperparameter optimization of deep learning models for wind power prediction', *Renewable Energy*, 221: 119700.

Hanifi, Shahram, Saeid Lotfian, Hossein Zare-Behtash, and Andrea Cammarano. 2022. 'Offshore Wind Power Forecasting—A New Hyperparameter Optimisation Algorithm for Deep Learning Models', *Energies*, 15: 6919.

Jagtap, Ameya Dilip, and George Em Karniadakis. 2022. 'How important are activation functions in regression and classification? A survey, performance comparison, and future directions', *ArXiv*, abs/2209.02681.
